# Diagnosing pediatric mitochondrial disease: lessons from 2,000 exomes

**DOI:** 10.1101/2021.06.21.21259171

**Authors:** Sarah L. Stenton, Masaru Shimura, Dorota Piekutowska-Abramczuk, Peter Freisinger, Felix Distelmaier, Johannes A. Mayr, Christine Makowski, Boriana Büchner, Bader Alhaddad, Charlotte L. Alston, Anna Ardissone, Rui Ban, Ivo Barić, Riccardo Berutti, Theresa Brunet, Elżbieta Ciara, Dasha Deen, Julien Gagneur, Daniele Ghezzi, Mirjana Gusic, Tobias B. Haack, Maja Hempel, Ralf A. Husain, Daniela Karall, Stefan Kölker, Urania Kotzaeridou, Thomas Klopstock, Robert Kopajtich, Vassiliki Konstantopoulou, Steffen Liez, Dominic Lenz, Albert Z. Lim, Hanna Mandel, Robert McFarland, Wolfgang Müller-Felber, Gerard Muñoz-Pujol, Akira Ohtake, Yasushi Okazaki, Rikke Olsen, Ewa Pronicka, Angela Pyle, Antonia Ribes, Dariusz Rokicki, René Santer, Manuel Schiff, Markus Schuelke, Dmitrii Smirnov, Wolfgang Sperl, Tim Strom, Frederic Tort, Polina Tsygankova, Rudy van Coster, Patrick Verloo, Jürgen-Christoph von Kleist-Retzow, Ekkehard Wilichowski, Tekla Wolstein, Manting Xu, Vicente Yépez, Michael Zech, Saskia Wortmann, Matias Wagner, Costanza Lamperti, Robert W. Taylor, Fang Fang, Agnés Rötig, Kei Murayama, Thomas Meitinger, Holger Prokisch

## Abstract

**Background:** The spectrum of mitochondrial disease is genetically and phenotypically diverse, resulting from pathogenic variants in over 400 genes, with aerobic energy metabolism defects as a common denominator. Such heterogeneity poses a significant challenge in making an accurate diagnosis, critical for precision medicine.

**Methods:** In an international collaboration initiated by the European Network for Mitochondrial Diseases (GENOMIT) we recruited 2,023 pediatric patients at 11 specialist referral centers between October 2010 and January 2021, accumulating exome sequencing and HPO-encoded phenotype data. An exome-wide search for variants in known and potential novel disease genes, complemented by functional studies, followed ACMG guidelines.

**Results:** 1,109 cases (55%) received a molecular diagnosis, of which one fifth have potential disease-modifying treatments (236/1,109, 21%). Functional studies enabled diagnostic uplift from 36% to 55% and discovery of 62 novel disease genes. Pathogenic variants were identified within genes encoding mitochondrial proteins or RNAs in 801 cases (72%), while, given extensive phenotype overlap, the remainder involved proteins targeted to other cellular compartments. To delineate genotype-phenotype associations, our data was complemented with registry and literature data to develop “GENOMITexplorer”, an open access resource detailing patient- (n=3,940), gene- (n=427), and variant-level (n=1,492) associations (prokischlab.github.io/GENOMITexplorer/).

**Conclusions:** Reaching a molecular diagnosis was essential for implementation of precision medicine and clinical trial eligibility, underlining the need for genome-wide screening given inability to accurately define mitochondrial diseases clinically. Key to diagnostic success were functional studies, encouraging early acquisition of patient-derived tissues and routine integration of high-throughput functional data to improve patient care by uplifting diagnostic rate.

## Introduction

Mitochondrial disease (MD) is an umbrella term for a group of complex metabolic disorders predominantly affecting mitochondrial oxidative phosphorylation (OXPHOS) (Gorman et al., 2016). To date, pathogenic variants in over 400 genes have been identified in both mitochondrial (mtDNA) and nuclear DNA, accounting for approximately 10% (416/4,022) of disease-associated genes in the Online Mendelian Inheritance in Man (OMIM) database (https://omim.org). MD has a minimum prevalence rate of 1 per 5,000 (Gorman et al., 2015). The absence of reliable biomarkers has led to MDs largely being defined by clinical criteria, some of which have been formalized into scoring systems (Morava et al., 2006, Witters et al., 2018). However, due to marked phenotypic overlap with non-MDs (Wortmann et al., 2015), here termed “MD-phenocopies”, limited phenotypic congruence, even between MD patients of shared molecular cause, and conversely, vast genetic heterogeneity amongst MD patients with a similar phenotype, MDs are most accurately defined on the molecular genetic level.

In recent years, genome-wide approaches including whole exome sequencing (WES) (Taylor et al., 2014, Ohtake et al., 2014, Wortmann et al., 2015, Legati et al., 2016, Kohda et al., 2016, Pronicka et al., 2016, Puusepp et al., 2018, Theunissen et al., 2018) and whole genome sequencing (WGS) (Riley et al., 2020) have been utilized in the diagnostic setting for MD. These approaches have overcome the need to prioritize candidate genes for targeted analysis in MD, such as by an invasive tissue biopsy followed by measurement of mitochondrial respiratory chain complex (RCC) activities (Wortmann et al., 2017). Current studies analyzing up to 142 patients from single centers report a wide range of WES/WGS diagnostic yields in suspected cases of MD (35-62%) depending on the age of onset, clinical evidence of MD, and the confirmation of a mitochondrial RCC defect (reviewed in Stenton and Prokisch 2020).

Here, analyzing over 2,000 patients by WES in an international multi-center collaborative effort, we uncover the genetic landscape of suspected MD, demonstrating high impact on clinical practice by identifying diagnoses with defect-specific therapy benefiting from early intervention and outlining a trajectory for the routine integration of high-throughput functional data to improve patient care by uplifting diagnostic rate.

## Methods

### Study population

We performed an analysis of 2,023 pediatric-onset (<18 years) patients analyzed by WES by one of 11 diagnostic referral centers specializing in MD in Europe and Asia between October 2010 and January 2021 (**Table S1**). Patients were eligible for inclusion if: (i) no genetic diagnosis was made by targeted sequencing (including complete mtDNA sequencing in some centers) or no prior genetic testing had been performed, and (ii) the patient had at least a “possible” MD according to the Nijmegen Mitochondrial Disease Criteria (MDC) (Morava et al., 2006). 293/2,023 patients (14%) were previously reported as part of focused research studies or novel disease gene descriptions (49 MD genes, 13 MD-phenocopy genes, **Table S2**). All participants or their legal guardians completed written informed consent and the study was approved by local ethical committees of the participating centers. A summary of the analysis workflow and study resources is depicted in **Fig. 1**.

**Figure 1.**
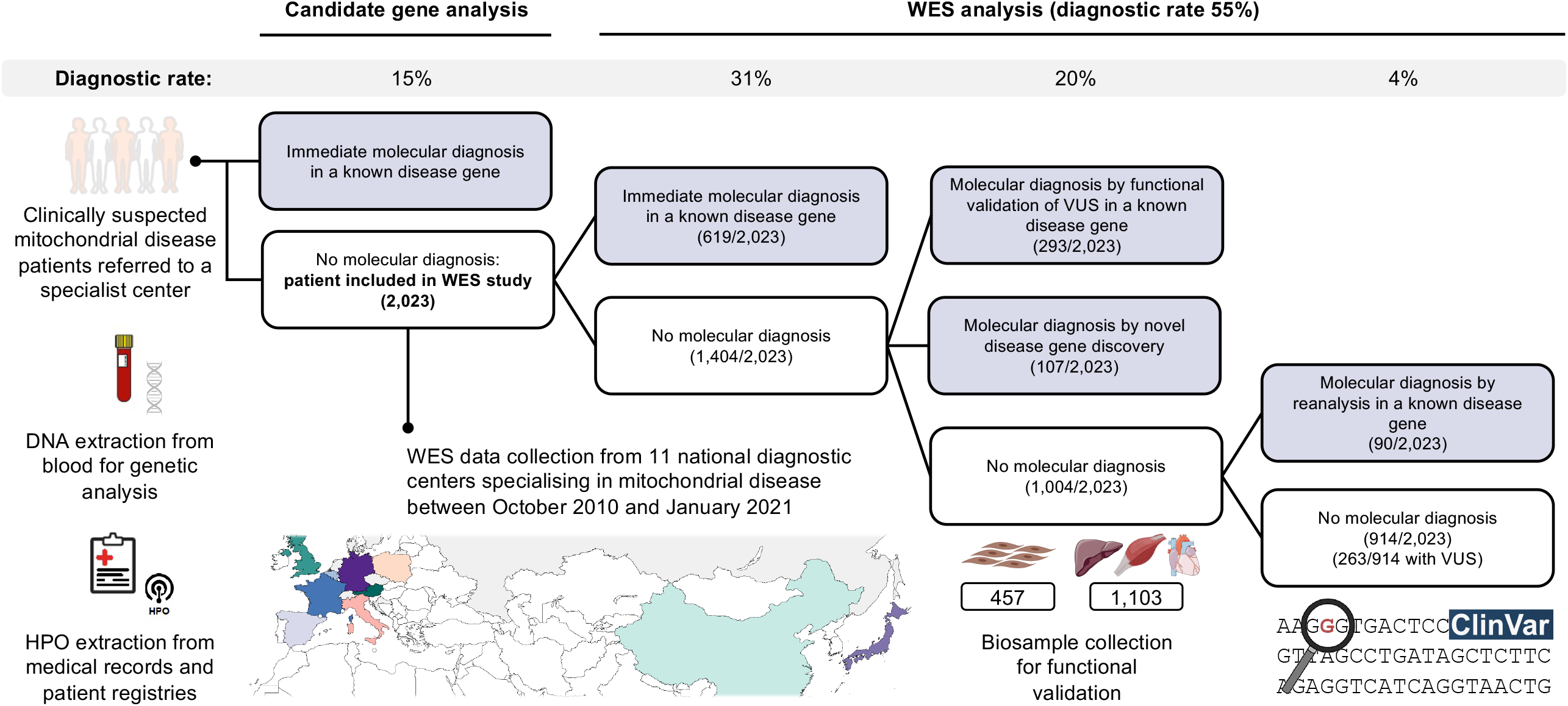
Diagnostic workflow for patients clinically suspected of MD. Based on the experience of the mitochondrial centers in the UK and Italy, an estimated 15% of pediatric patients receive a molecular diagnosis by targeted gene sequencing prior to WES (≤5 individual genes or mtDNA sequencing), the majority of which have pathogenic variants in the mtDNA. This study analyzed 2,023 patients investigated by WES. Patients were referred to one of 11 diagnostic referral centers specialized in MD (**Table S1**). DNA was extracted from blood for WES and clinical data were collected from the medical records and/or patient registries for HPO extraction. Patient DNA was sequenced between October 2010 and January 2021. The WES diagnostic rate was stratified by immediate diagnoses (i.e., the identification of ACMG classified “pathogenic” or “likely pathogenic” variants in a known disease gene in-keeping with the patient’s clinical presentation) (619/2,023, 31%), diagnoses in known disease genes requiring further functional studies (293/2,023, 14%), diagnoses leading to the discovery and functional validation of a novel disease gene (107/2,023, 5%, 49 novel MD genes and 13 novel MD-phenocopy genes), and diagnoses identified by reanalysis of the WES data (90/2,023, 4%, mean 2.6 year ± 1.7 year s.d. interval required). The cases solved by reanalysis were retrospectively identified as those with “pathogenic” or “likely pathogenic” variants in disease genes only described to be disease-associated following the initial WES analysis. Of these cases, 53 molecular diagnoses were made in MD genes and 37 in MD-phenocopy genes. The resources for functional validation studies included 457 patient-derived fibroblast cell lines (23% of the cohort) and 1,103 tissue biopsy samples including skeletal muscle, cardiac muscle, and liver (55% of the cohort). HPO, human phenotype ontology; WES, whole exome sequencing.

### Genetic analyses

Experimental procedures were performed in the respective centers, as detailed in the **Supplemental Methods**. All disease-causing variants were classified according to the ACMG criteria (Richards et al., 2015) (**Table S3**) and were submitted to the ClinVar database or are in progress.

### Mitochondrial disease gene definition

MD genes were defined as per Schlieben and Prokisch 2020 (n=413), with the addition of three recently identified MD genes (*DNAJC30, LIG3*, and *MRPL38*) (Stenton et al., 2021, Kopajtich et al., 2021, Bonora et al., 2021). This definition included 49 of 62 novel gene discoveries made in this cohort (**Table S2**).

### Phenotype analyses

Patient phenotype data were extracted as human phenotype ontology (HPO) terms (Köhler et al., 2021) from medical records and/or patient registries. All ancestral HPO terms were derived from the ontology using the R package “OntologyX” (Greene et al., 2017). MDC scoring was standardized for the usage of HPO terms (**Supplemental Methods**). Symmetric semantic similarity (SS) was calculated between patient HPO terms and established disease-gene associated HPO terms using the R package “PCAN” and the HPO “genes to phenotype” annotation files (human-phenotype-ontology.github.io/downloads.html, accessed April 2021), limited to disease genes in the OMIM database (https://www.omim.org, accessed April 2021) (n=4,022).

### Statistical analyses

Statistical analyses were performed in R version 3.6.1. Enrichment analyses used the Fisher’s exact test. Bonferroni correction was used to adjust p values for multiple testing. A significance level of 0.05 was used. Correlation was tested using Spearman’s rank method. Diagnostic test performance was analyzed by ROC (receiver operating characteristic) curves and calculation of the AUC (area under the ROC curve) with 95% confidence intervals (CI).

## Results

### Study population description

2,023 pediatric patients (1,950 index patients plus 73 affected siblings) were analyzed by singleton (1,779/2,023, 88%) or trio (244/2,023, 12%) WES. 1,066/2,023 patients were male (53%). The age of onset ranged from neonatal to juvenile, with over one third of patients presenting in infancy (709/2,023, 35%) (**Table 1**).

**Table 1.** Key demographic, genetic, clinical, and biochemical features stratified by molecular diagnosis.

|  |  | **Total** | **MD** | **MD-phenocopy** | **Unsolved** |
| --- | --- | --- | --- | --- | --- |
| **Patients** |  | 2,023 | 801/2,023 (40%) | 308/2,023 (15%) | 914/2,023 (45%) |
| **Gender** | Male | 1,066/2,023 (53%) |  |  |  |
| **Onset** | Neonatal<br>(≤28 days) | 545/2,023 (27%) |  |  |  |
|  | Infantile<br>(>28 days, <2 years) | 709/2,023 (35%) |  |  |  |
|  | Early childhood<br>(≥2 years, <5 years) | 444/2,023 (22%) |  |  |  |
|  | Childhood<br>(≥5 years, <12 years) | 217/2,023 (11%) |  |  |  |
|  | Juvenile<br>(≥12 years, <18 years) | 107/2,023 (5%) |  |  |  |
| **Diagnostic rate** | Study population | 1,109/2,023 (55%),<br>94/1,109, 8% *de novo* | 801/2,023 (40%),<br>33/801, 4% *de novo* | 308/2,023 (15%),<br>61/308, 20% *de novo* |  |
|  | Singleton WES,<br>*de novo* | 986/1,779 (55%),<br>60/986, 6% *de novo* |  |  |  |
|  | Trio WES,<br>*de novo* | 123/244 (50%),<br>34/123, 28% *de novo* |  |  |  |
| **Median HPO**<br>**phenotypes (range)** |  | 8 (2-40) | 8 (2-40) | 7 (2-31) | 8 (2-35) |
| **Median systems involved (range)** |  | 4 (1-12) | 4 (1-12) | 4 (1-11) | 4 (1-11) |
| **Abnormal RCC activity** |  | 889 (44%) | 399 (50%) | 104 (34%) | 386 (42%) |
| **Elevated lactate (serum and/or CSF)** |  | 1,024 (51%) | 509 (64%) | 97 (31%) | 418 (46%) |
| **MRI abnormality (BG and/or BS)** |  | 524 (26%) | 270 (34%) | 45 (15%) | 209 (23%) |
| **MDC score** | Nijmegen MDC score | 6.0 ± 2.3 | 6.7 ± 2.4 | 5.2 ± 2.2 | 5.8 ± 2.2 |
|  | Revised MDC score | 4.9 ± 2.0 | 5.4 ± 2.1 | 4.5 ± 1.8 | 4.7 ± 2.0 |

WES, whole exome sequencing; HPO, human phenotype ontology; RCC, respiratory chain complex; CSF, cerebral spinal fluid; MRI, magnetic resonance imaging; BG, basal ganglia; BS, brain stem; MDC, mitochondrial disease criteria.

In total, 949 unique non-redundant HPO terms were extracted, a median of eight HPO terms per patient (range 2-40) across a median of four organ systems (range 1-12) (**Table 1, Supplemental Fig. 1a-b**). 321/2,023 patients (16%) presented with a constellation of symptoms indicative of Leigh syndrome (**Table S4**, Rahman et al., 1996) and only 37/2,023 patients (2%) with symptoms suggestive of MELAS syndrome (Mitochondrial Encephalopathy, Lactic Acidosis, and Stroke-like episodes, n=20), Alpers syndrome (n=10), Kearns-Sayre syndrome (n=2), MERRF (Myoclonic Epilepsy with Ragged Red Fibers, n=1), NARP (Neuropathy, Ataxia, and Retinitis Pigmentosa, n=1), and LHON (Leber Hereditary Optic Neuropathy, n=3). The remaining 1,665/2,023 patients (82%) did not present with a distinct MD syndrome.

### Diagnostic rate

A molecular diagnosis was made by WES in 1,109/2,023 patients (55%) (**Fig. 1** and **Fig. 2a**). A dual diagnosis was identified in 7 patients (0.3%). In 263/2,023 patients (13%), the WES analysis remained unsolved with variants of uncertain significance (VUS) identified in OMIM disease genes or candidate disease genes. In the remaining 651/2,023 patients (32%) no variant could be prioritized for further consideration. By comparison, applying a filter restricting the observed disease-causing variation to genes covered by 13 “mitochondrial disease” panels (see **Supplemental Methods**) resulted in a diagnostic rate of 27% (± 7% s.d.) (**Supplemental Fig. 2a**).

**Figure 2.**
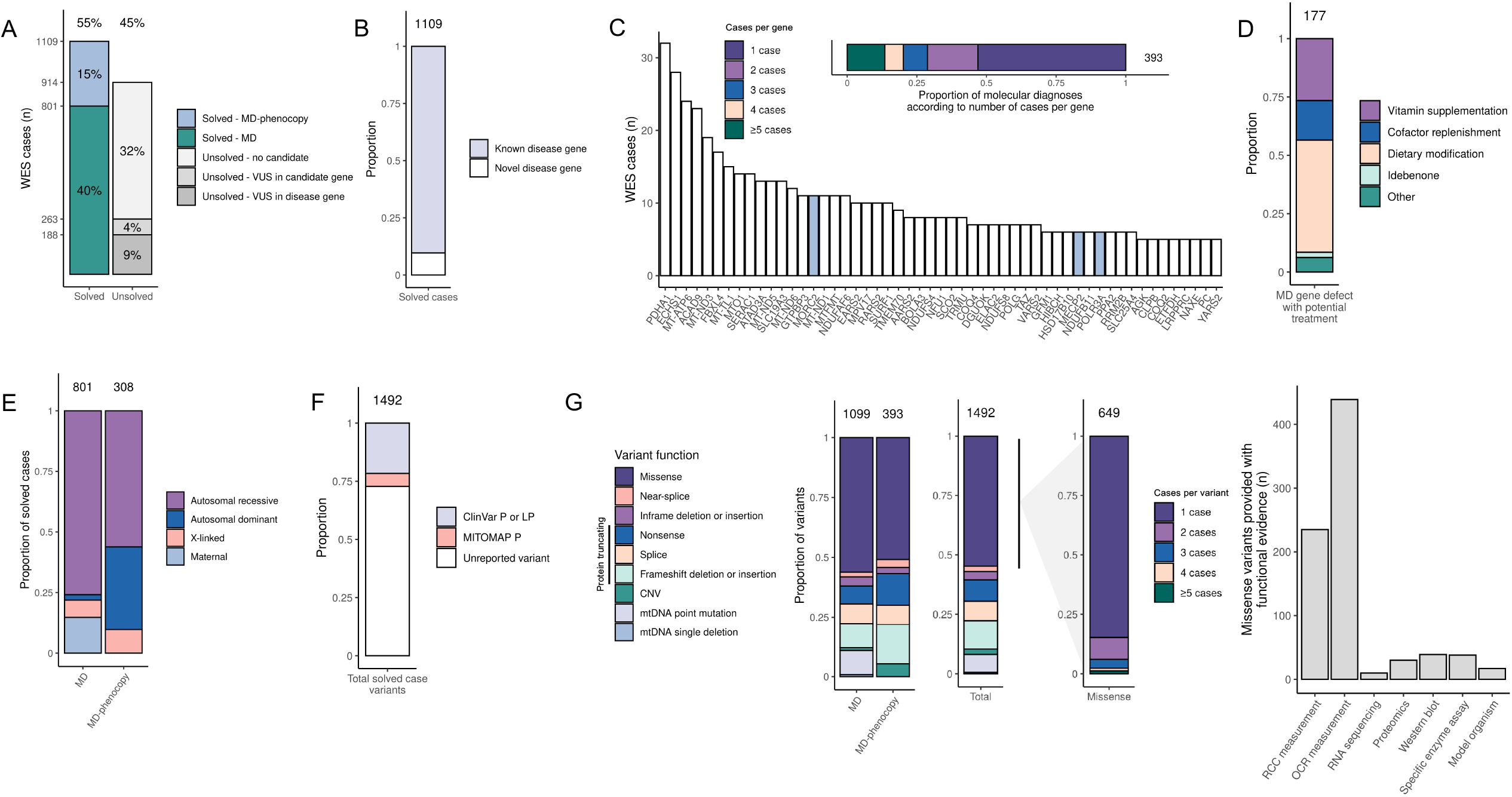
Molecular etiology of the MD and MD-phenocopies. **A**, Number of solved and unsolved cases stratified by underlying molecular cause. **B**, Proportion of solved cases in known and novel disease genes (defined as novel at the point of molecular diagnosis, i.e., whether the case was involved in one of the 62 novel disease gene discoveries). **C**, Proportion of molecular diagnoses made in a single case and multiple cases. Those detected in ≥5 cases are individually displayed with their frequency. The most frequent genes with disease-causing variants in the MD-phenocopy patients are highlighted in light blue. **D**, Solved MD patients with variants in one of 29 MD genes reported to have a potential treatment option, stratified by treatment type. **E**, Proportion of molecular diagnoses by mode of inheritance in the MD and MD-phenocopy patients. **F**, Proportion of variants listed in ClinVar or MITOMAP as “pathogenic” (P) or “likely pathogenic” (LP) at the point of molecular diagnosis. **G**, Proportion of variants of each functional class in the MD patients, MD-phenocopy patients, and all solved cases collectively. Most variants were missense. The proportion of missense variants detected in a single case and multiple cases is depicted with the corresponding functional validation study leading to designation of the variant as “pathogenic” or “likely pathogenic”. WES, whole exome sequencing; VUS, variant of uncertain significance; CNV, copy number variant.

### Molecular genetic etiologies

The disease-causing variants were detected in 199 different MD genes (150 known and 49 novel) in 801/1,109 (72%) solved cases (**Fig. 2a-b**). In the remaining 308/1,109 solved cases (28%), the molecular diagnosis involved a MD-phenocopy gene (181 known and 13 novel disease genes) primarily implicated in neurodevelopmental disease, neuromuscular disease, or other inborn errors of metabolism (**Supplemental Fig. 2b**).

The molecular diagnoses spanned 393 genes, many of which were reported in single cases only (208/393, 53%). The most frequent MD gene hits were *PDHA1, ECHS1, MT-ATP6*, and *ACAD9* each with ≥23 cases (**Fig. 2c**). Amongst the MD gene hits were 29 defects identified in 177 patients where defect-specific vitamin supplementation, replenishment of a critical cofactor, dietary modification, or idebenone therapy offers the potential for targeted disease-modifying treatment (**Fig. 2d**) (**Table S5**) (Bick et al., 2020, Hoytema van Konijnenburg et al., 2021, Distelmaier et al., 2017). The most frequent MD-phenocopy gene hits were *MORC2, MECP2*, and *POLR3A*, each with ≥6 cases.

Pathogenic mtDNA variants accounted for 15% (118/801) of MD molecular diagnoses and were mostly heteroplasmic (71/118 cases, 60%). In the remaining majority (683/801, 85%) the causative gene defect was nuclear-encoded and inherited in an autosomal recessive manner (606/683 cases, 89%) (**Fig. 2e**), of which 306/606 harbored a homozygous causative variant (51%). When searching for runs of homozygosity, a first- or second-degree parental relationship was suspected in a total of 44/2,023 cases, accounting for 35/306 cases with homozygous “pathogenic” or “likely pathogenic” variants in MD genes (11%). In contrast to MD, *de novo* dominant variants were responsible for disease in significantly more MD-phenocopy cases (4% of MD and 20% of MD-phenocopy, p-value 1.5×10^−15^, Fisher’s exact test) (**Table 1**). However, *de novo* variants were also frequently detected in mtDNA-encoded MD genes (7 cases) and in the nuclear-encoded MD gene *PDHA1* (14 cases, four by trio analysis and 10 by detection of the heterozygous variant in singleton analysis followed by parental sequencing).

In total, 1,492 disease-causing variants were reported (1,173 unique variants) (**Table S3, Supplemental Fig. 2c-e**). 406/1,492 variants (27%) were already documented as “pathogenic” or “likely pathogenic” in ClinVar or MITOMAP at the time the patient’s molecular diagnosis was made (**Fig. 2f**). Missense variants accounted for the largest proportion of all disease-causing variants (817/1,492 variants, 55%), of which the majority (550/649 unique missense variants, 85%) were reported in single cases only. In 304/538 cases carrying a missense variant (57%), the variant was homozygous or compound heterozygous with a second missense variant, posing a challenge to interpretation of pathogenicity and requiring functional studies to provide evidence of pathogenicity (**Fig. 1** and **Fig. 2g**). In contrast, biallelic null variants were identified in just 162/1,109 solved cases (15%). Notably, in 33/1,109 solved cases, pathogenic CNVs were identified, of which in three cases, the CNV was in *trans* with a single nucleotide variant.

### Genotype-phenotype associations

Most patients came to medical attention with neurological (1,568/2,023, 78%), metabolic (1,541/2,023, 76%), muscular (1,066/2,023, 53%), digestive (579/2,023, 29%), and/or cardiological (574/2,023, 28%) symptoms (**Fig. 3a** and **Supplemental Fig. 3a**). In solved MD patients, the most frequently reported clinical and imaging findings were neurodevelopmental abnormality (546/801, 68%) and abnormality of basal ganglia and/or brainstem MRI signal intensity (404/801, 50%). The most prevalent laboratory findings were increased serum lactate (506/801, 63%) and abnormal activity of mitochondrial RCC enzymes (389/801, 49%) (**Fig. 3a**). Across the entire cohort, 83 HPO terms were reported in ≥50/2,023 patients. None of these HPO terms were entirely specific for MD. However, 9/83 HPO terms were significantly enriched in the solved MD patients in comparison to the MD-phenocopies, including HPO terms indicative of Leigh-like syndrome (encephalopathy, abnormality of the basal ganglia and/or brainstem, and increased serum lactate), mitochondrial RCCI defect, and cardiomyopathy (**Fig. 3a, Table S6**). In combination, HPO terms indicative of Leigh syndrome (**Table S4**) were highly suggestive of MD (OR 3.25, 95% CI 2.0- 5.5, p 1.1×10^−7^, Fisher’s exact test).

**Figure 3.**
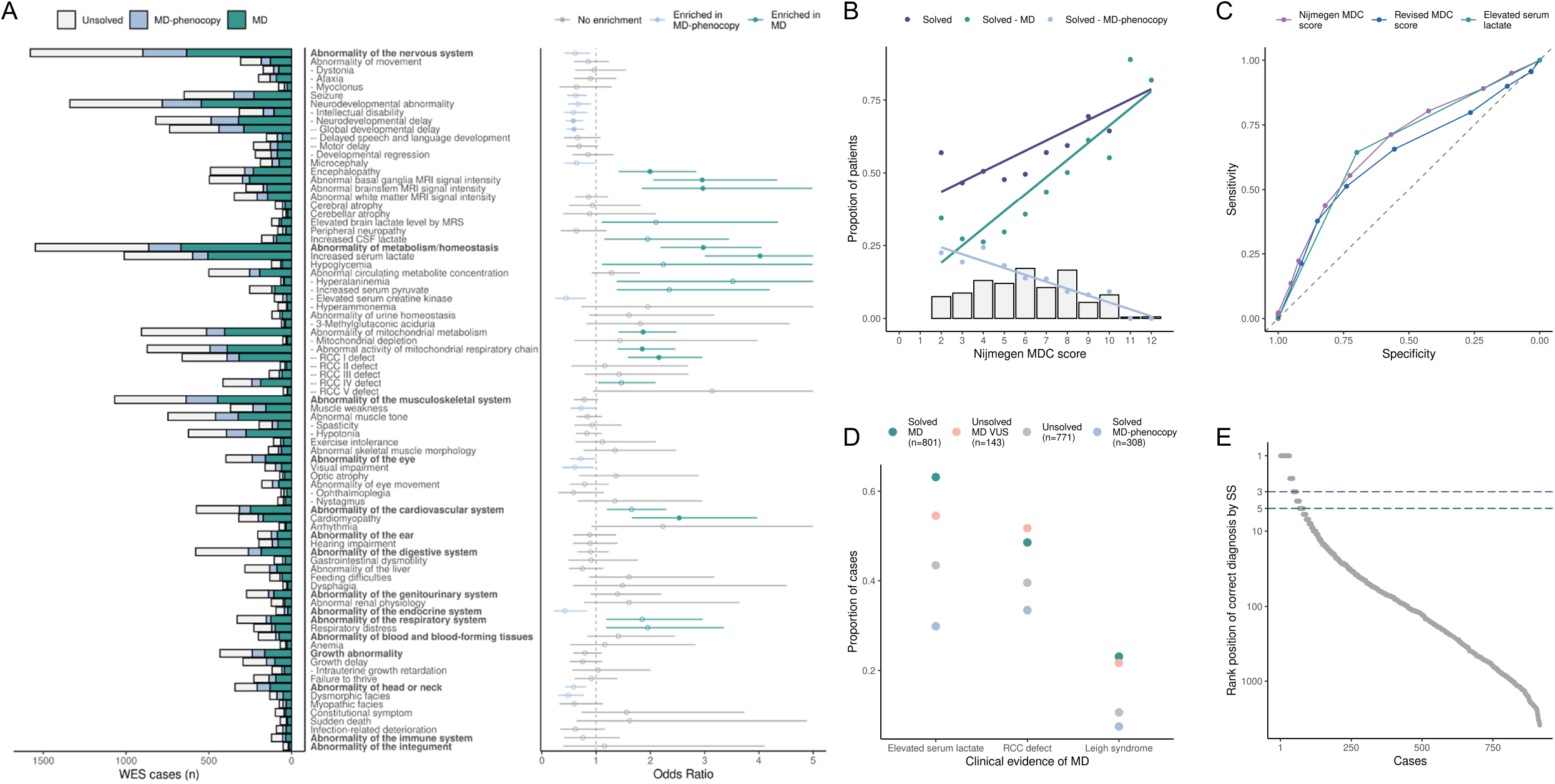
Comprehensive overview of patient phenotype and association with the molecular diagnosis. **A**, Frequency of HPO terms reported in ≥50/2,023 patients, stratified by the underlying molecular diagnosis (left panel). HPO term enrichment analysis comparing solved MD and MD-phenocopy patients (right panel) depicting the odds ratio with the 95% confidence interval (confidence interval limits >5 are not shown, see **Table S6**). Nominally significant results are depicted in color and results with multiple testing corrected significance are depicted with filled shapes (adjusted p-value ≤0.05). **B**, Distribution of Nijmegen MDC score (grey bars) in the study population and the corresponding correlation to the diagnostic rate (purple), proportion of mutations in MD genes (green), and proportion of mutation in MD-phenocopy genes (blue). Increase in the Nijmegen MDC score was reflected by increase in the overall diagnostic rate (R 0.86, p 1.3×10^−3^, Spearman’s rank correlation), increase in the proportion of solved MD (R 0.92, p 2.2×10^−16^, Spearman’s rank correlation), and decrease in the proportion of MD-phenocopies (R -0.96, p 2.4×10^−6^, Spearman’s rank correlation). The unexpectedly high diagnostic rate and proportion of solved MD at a Nijmegen MDC score of 2 (120 cases) reflects 114 legacy cases where only limited clinical information was available and six cases with single organ involvement (e.g., optic atrophy). **C**, Receiver operator characteristic (ROC) curves for the Nijmegen MDC score (AUC MD vs. MD-phenocopy 0.68, 95% CI 0.64-0.71), Revised MDC score (AUC MD vs. MD-phenocopy 0.63, 95% CI 0.60-0.67), and elevated serum lactate (AUC MD vs. MD-phenocopy 0.67, 95% CI 0.64-0.70) in stratifying the MD and MD-phenocopy patients. The Revised MDC score was included as an alternative MDC, as in contrast to the Nijmegen MDC score the Revised MDC score does not consider tissue histopathology and tissue biochemistry findings (see **Supplemental methods**). **D**, Proportion of patients meeting criteria indicative of MD in our study population, stratified by underlying molecular diagnosis. **E**, Solved patients ordered by the rank of their respective disease-causing gene amongst all OMIM disease genes by HPO phenotype semantic similarity (SS). Patients above the horizontal lines mark those in whom the correct gene defect was predicted by HPO phenotype within the highest three and five SS scores, respectively. WES, whole exome sequencing; MDC, mitochondrial disease criteria; AUC, area under the curve; BG, basal ganglia; BS, brain stem; RCC, respiratory chain complex; SS, semantic similarity.

To objectively categorize patients by the level of clinical evidence of MD, we applied the Nijmegen MDC score (Morava et al., 2006), demonstrating a continuum from “possible” (590/2,023, 29%, score 2-4) to “probable” (804/2,023, 40%, score 5-7) and “definite” (629/2,023, 31%, score 8-12) MD (**Fig. 3b**). Increase in the Nijmegen MDC score was reflected by increase in the overall diagnostic rate, increase in the proportion of molecularly diagnosed MD, and decrease in the proportion of MD-phenocopies. Notably, a “definite” MD (score 8-12) was reported in 55/308 MD-phenocopy patients. Given these data, no clear threshold of Nijmegen MDC score for the accurate diagnosis of an MD can be drawn in our cohort (**Fig. 3b**) and the ability of both Nijmegen and Revised MDC (Witters et al., 2018) scoring and elevated serum lactate was found to be limited in discerning between MD and MD-phenocopy (**Fig. 3c, Table S7**).

In 188/914 (21%) unsolved cases, VUS were detected in known disease genes in keeping with the expected mode of inheritance. In a subset (143/188, 76%), the VUS occurred in one of 72 MD genes or 43 candidate disease genes with strong evidence of mitochondrial localization (according to Mitocarta3.0). In comparison to the remaining unsolved cases, these 143 cases had significantly more reports of elevated serum lactate, mitochondrial RCC defect(s), and a phenotype combination indicative of Leigh syndrome (**Fig. 3d, Table S8**), clinical features providing supportive evidence for the pathogenicity of the variant(s). However, current unavailability of fibroblast cell lines hindered further functional studies to confirm pathogenicity, highlighting the importance of acquiring these bio-samples early in the diagnostic process.

Akin to MDC scoring, the ability of HPO semantic similarity (SS) scoring to retrospectively predict the patient’s molecular diagnosis was limited. The correct molecular diagnosis had the highest SS score amongst all OMIM disease genes in just 3% of cases, increasing to 6% and 9% when considering the three and five highest SS scores, respectively (**Fig. 3e**). Overall, more complex phenotypes correlated with improved SS scoring (R 0.39, p 2.2⨯10^−16^, Spearman’s rank correlation) (**Supplemental Fig. 3b**).

### GENOMITexplorer

The open access online resource “GENOMITexplorer” lists all “pathogenic” and “likely pathogenic” variants reported in this study with the respective predicted function, heteroplasmy level, allele frequency (according to gnomAD), CADD score, SIFT score, ACMG classification, functional evidence, and phenotype SS score. HPO-gene associations for all patients included in the study (n=2,023) in addition to pediatric molecularly diagnosed patients from mitochondrial disease registries (mitoNET and Besta, n=318) and from literature reports (n=1,599) are provided on the patient- (n=3,940, 3,026 molecularly diagnosed), gene- (n=427), and variant level (available for n=1,492). The most discriminating phenotypes by genetic diagnosis can be explored for all genes with ≥5 (n=114) reported patients carrying “pathogenic” and “likely pathogenic” variants.

## Discussion

We analyzed over 2,000 pediatric patients by WES under the clinical suspicion of MD. This is the largest study of its kind and was made possible only by broad international collaboration. A molecular diagnosis was made in just over 50% of cases and 62 novel disease genes discoveries were made in the cohort over a 10-year period (49 MD, 13 MD-phenocopy), illuminating underlying disease mechanism for the application or development of specific treatments not considered previously. This high diagnostic rate was achieved by close interconnection between research and routine diagnostics, and places suspected MD amongst the highest yielding metabolic and neurological primary indications for WES (on average 20-35%) (**Supplemental Fig. 4a**).

By nature, the study focused on more difficult cases referred to specialist centers. Therefore, despite providing a reliable estimate of the power of WES in MD given the size of the study, it does not intend to provide an epidemiological perspective. The diagnostic rate is conservative, as across our centers we diagnose at least 15% of patients without WES, mostly by mtDNA sequencing (see **Fig. 1**). Further, we do not see the patients diagnosed prior to referral to a specialist center. Therefore, had WES been applied as a first step in the diagnostic pathway in all patients, the yield would have been considerably higher. This is reflected by the most recent 250 patients analyzed in Munich, Germany, where WES is the first genetic test in the majority and the diagnostic rate was >60%. Moreover, in a further 7% (143/2,023) of patients in the study, we identified VUS in established or candidate MD genes with clear clinical evidence of MD and thereby high likelihood for pathogenic designation upon future functional studies.

Functional studies of patient-derived material provided significant value for variant interpretation, necessitated by the preponderance of private missense variants with no prior functional annotation (>80%). Without functional validation, our diagnostic rate would have been capped at approximately 35% (see **Fig. 1**), strongly arguing for proactive bio-sampling of patients, (e.g., a minimally invasive skin biopsy to establish a fibroblast cell line), and parallel analysis of WES and functional data. Fibroblasts are an excellent resource for the study of many MDs, as they may be used for mitochondrial RCC measurement with results comparable to invasive muscle biopsy (Ogawa et al., 2017, Wortmann et al., 2017) and detection of impaired mitochondrial respiration by measurement of oxygen consumption rate or other biomarkers. Moreover, fibroblasts open the option for multi-omic studies, pioneered in MD, such as RNA sequencing and proteomics (Kremer et al., 2017, Kopajtich et al., 2021, Yepez et al., 2021). These multi-omics assays pave the way for integrated high throughput functional readouts of variant consequence, allowing interpretation of the vast number of VUS detected by WES/WGS.

With the continuous discovery of novel disease genes (>300 per year in OMIM and 212 MD genes during the course of this study) (**Supplemental Fig. 4b**), the expansion of genotype-phenotype associations, and the description of different modes of inheritance for established disease genes, WES reanalysis becomes an essential component of the diagnostic pathway. Reanalysis uplifted our diagnostic rate by almost 5% (see **Fig. 1**). A prime example was the diagnosis of 11 patients with pathogenic *de novo* dominant variants in *MORC2* by reanalysis following a report expanding the phenotypic spectrum to include neurodevelopmental and metabolic abnormalities (Guillen Sacoto et al., 2020).

Reaching a genetic diagnosis is important for defect-specific treatments, disease prevention, and genetic and reproductive counselling. Moreover, it is a prerequisite for inclusion in constantly ongoing clinical trials, an area in which there has been immense progress driven by increased understanding of disease pathomechanism and the unmet clinical need, reflected by over 50 registered clinical trials for MD (Russell et al., 2020). In our cohort, disease-modifying treatments, such as a ketogenic diet in *PDHA1* defects, are reported based on case reports for 29 of the identified MD gene defects, across 177 patients in our study (Bick et al., 2020, Hoytema van Konijnenburg et al., 2021, Distelmaier et al., 2017), as exemplified in our earlier publications (Stenton et al., 2021, Zhou. L. et al., 2020, Zhou. J. et al., 2020, Repp et al., 2018, Distelmaier et al., 2013, Haack et al., 2012). In addition, 43 of the identified disease genes in the MD- phenocopies across 61 patients, have reported treatments. In total, these 238 patients (236/1,109 solved cases, 21%) endorse an unbiased exome-wide approach to identify and optimize care for unexpected diseases without delay.

Despite the clinical challenges to diagnose MD correctly, over 70% of the solved cases were confirmed as such, with defects equally distributed across all classes of mitochondrial function (**Fig. 4**). A preponderance of nuclear DNA variants (85%) and autosomal recessive inheritance (>75%) in the solved MD patients was anticipated for a pediatric population (Barca et al., 2020). These figures are, however, inflated by ascertainment bias given thorough mtDNA analysis undertaken earlier in the diagnostic pathway in some centers. *PDHA1, ECHS1*, and *ACAD9* were the most frequent nuclear-encoded MD genes with pathogenic variants, in-keeping with large collections of patients in the literature (Lissens et al., 2000, Marti-Sanchez et al., 2020, Repp et al., 2018) and the report of founder mutations in *ECHS1* (Haack et al., 2015, Tetreault et al., 2015), encouraging the implementation of clinical trials for these defects. Nevertheless, certain mtDNA-encoded genes with wide clinical variability, such as *MT-ATP6*, were frequent hits in line with large mutation reports (Ganetzky et al., 2019, Ng et al., 2019, Stendel et al., 2020). This reinforces the value of calling mtDNA variants from WES data in a blood sample from children. The low frequency of certain nuclear-encoded molecular diagnoses in our study may be distorted by prior targeted sequencing of candidate disease genes, such as *POLG* (7 diagnoses) and *SURF1* (10 diagnoses), two of the most frequently reported nuclear-encoded MDs in the literature (Rahman and Copeland 2019, Barca et al., 2020, Tan et al., 2020). Importantly, >20% of genetic diagnoses were made in MD-phenocopy genes, only identifiable by opting for unbiased exome-wide capture of variation. Amongst the MD-phenocopies we report 6-fold more autosomal dominant and *de novo* dominant inheritance of disease, highlighting the importance of trio WES to streamline variant interpretation.

**Figure 4.**
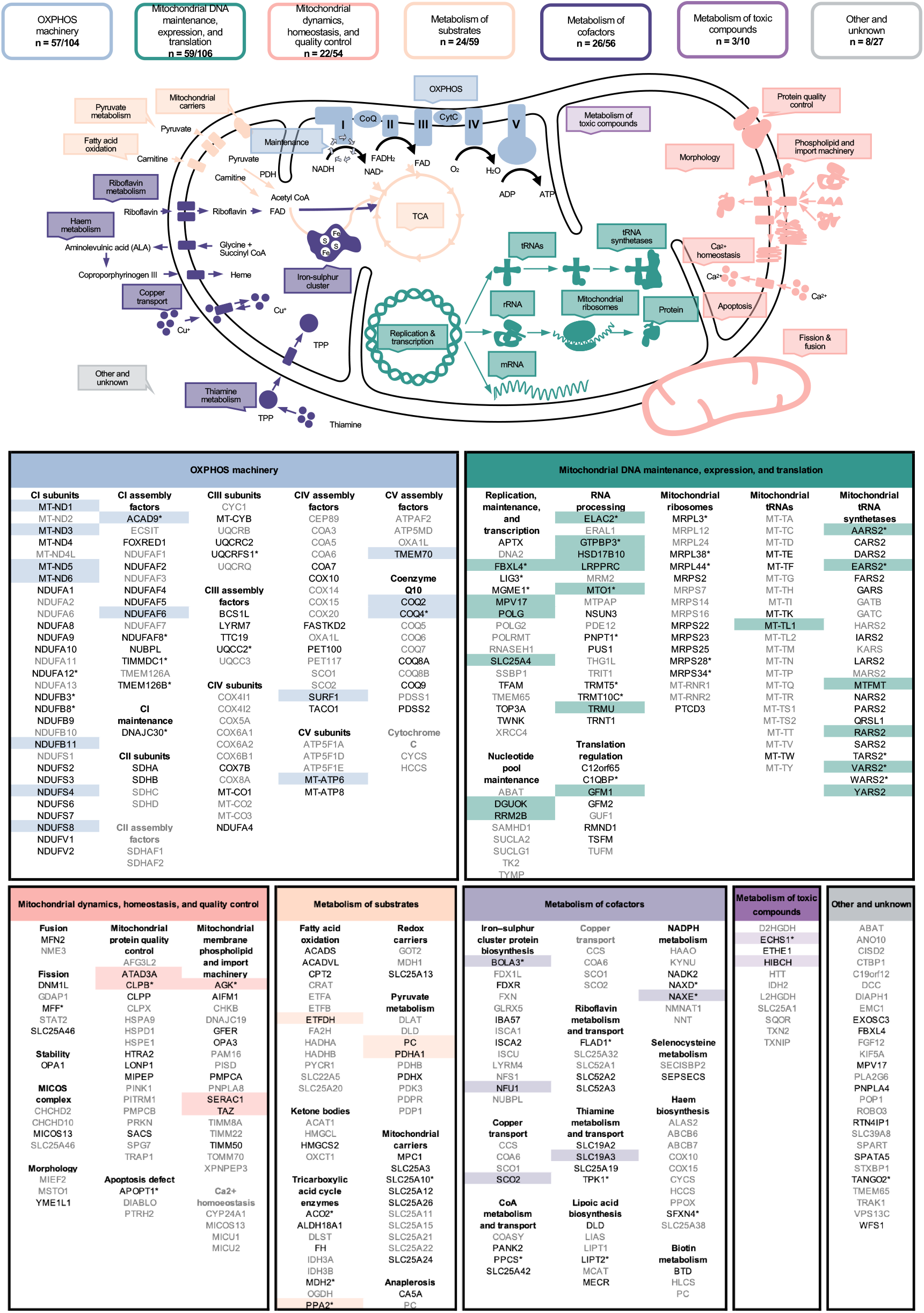
MD gene overview and variant detection in this study. Depiction of the functional category of all defined MD genes indicating the number of disease genes per functional category with disease-causing variants detected in this study. All disease genes are listed beneath. “Pathogenic” or “likely pathogenic” variants were detected in genes highlighted in color in ≥5 cases and in those depicted in black in ≥1 case. “Pathogenic” or “likely pathogenic” variants were not detected in the genes depicted in grey in this study. In total 199 of the 416 defined MD genes were reported with defects. Genes marked with an asterisk were novel disease gene discoveries made within the cohort over the 10-year period of this study (**Table S2**).

Within our study, encephalopathy, cardiomyopathy, MRI signal intensity changes in the basal ganglia and/or brainstem, elevated serum lactate, and mitochondrial RCC defects were found to be the most enriched HPO terms in molecularly diagnosed MD patients. However, given extensive phenotypic overlap with other Mendelian diseases, MDC scoring were unable to reliably stratify the MD and MD-phenocopy patients, and HPO phenotype semantic similarity scoring prioritized the correct molecular diagnosis in less than 5% of patients, a figure considerably lower than reported for general pediatric genetic studies (French et al., 2019). For these reasons, MD is most efficiently and reliably diagnosed by WES early in the diagnostic pathway to direct the need for specific clinical investigations and to prevent unnecessary interventions in children. Moreover, the clinical heterogeneity is not only within the spectrum of MD, but also between variants within a single gene defect (Marti-Sanchez et al., 2020), indicating the need to study MDs on the variant level. A starting point is GENOMITexplorer, providing patient- and variant-level resolution of genotype-phenotype associations, so far of only limited availability in the established databases (Firth et al., 2009).

We were unable to provide a molecular diagnosis for just under half of the investigated patients. The possible reasons for this are manifold: (i) due to pitfalls in variant prioritization, such as single heterozygous *de novo* variants, given the majority of patients underwent singleton WES analysis, and pitfalls in variant interpretation (e.g., hypomorphic and synonymous variants causing abnormal splicing, and deep intronic and regulatory variants), a limitation beginning to be addressed by multi-omic integration (Kopajtich et al., 2021); (ii) due to certain genetic variants being undetectable by WES given insufficient depth of coverage, locus-specific features (e.g., GC-rich regions and homopolymeric repeats), sequencing biases, and genomic alterations (e.g., large deletions, insertions, chromosomal rearrangements, short tandem repeats), the inability to delineate some of these alterations may be overcome by WGS, specifically long-read sequencing; (iii) due to the high number of VUS found in genes clinically compatible with the patient’s phenotype, the possibility to perform functional studies is fundamental to achieve a definitive diagnosis; and (iv) due to individuals with suspected MD carrying pathogenic variants in as yet unidentified disease-relevant genes, the discovery of which continues year-on-year (**Supplemental Fig. 4b**). Such disease gene discoveries, as exemplified by the 62 novel disease gene discovered in this cohort, are accelerated by collation of patient datasets to increase the signal for detection. In this manner, more challenging diagnoses can be made, such as variants with incomplete penetrance as recently described for *DNAJC30*, a novel recessive MD manifesting as LHON that is responsive to idebenone therapy (Stenton et al., 2021). This underpinned our motivation to form an international MD network and encourages patient referral to centers of expertise in which research projects flank state of the art diagnostics and act as recruitment centers for clinical trials.

Overall, our study demonstrates the power and efficiency of WES and functional validation in specialized centers in diagnosing patients with suspected MD, revealing that a substantial proportion of MD patients benefit from early diagnosis and targeted treatment, and exemplifying health economic benefits from the application of WES early in the diagnostic pathway.

### Online resources

GENOMITexplorer (prokischlab.github.io/GENOMITexplorer/)

GENOMIT (genomit.eu)

ClinVar (ncbi.nlm.nih.gov/clinvar/)

MITOMAP (mitomap.org)

gnomAD (gnomad.broadinstitute.org)

OMIM (omim.org)

## Data Availability

All data needed for the analysis presented in the manuscript are available on GENOMITexplorer (https://prokischlab.github.io/GENOMITexplorer/).

## Acknowledgements

The authors acknowledge support by the German Federal Ministry of Education and Research (BMBF, Bonn, Germany) awarded grant to the German Network for Mitochondrial Disorders (mitoNET, 01GM1906A), the German BMBF and Horizon2020 through the E-Rare project GENOMIT (01GM1920A, genomit.eu), and the ERA PerMed project PerMiM (01KU2016A, permim.eu). AR and MS acknowledge the Association Français contre les Myopathies, GIS-Institut des Maladies Rares, French Agence Nationale pour la Recherche (A.N.R.), Association contre les Maladies Mitochondriales (AMMi), Fondation Maladies Rares, Collaboration CEA/IG/CNG-INSERM, MSD-Avenir for Devo-Decode project, and Bioresources of the Necker Imagine DNA biobank (BB-033-00065). RM and RWT are supported by the Wellcome Centre for Mitochondrial Research (203105/Z/16/Z), the Medical Research Council (MRC) International Centre for Genomic Medicine in Neuromuscular Disease (MR/S005021/1), the Mitochondrial Disease Patient Cohort (UK) (G0800674), the Lily Foundation and the UK NHS Specialised Commissioners who fund the “Rare Mitochondrial Disorders of Adults and Children” Service in Newcastle upon Tyne. CLA is supported by an NIHR postdoctoral fellowship (PDF- 2018-11-ST2-02). AR, GMP, and FT were supported by the Instituto de Salud Carlos III (PI16/01048; PI19/01310) (Co-funded by European Regional Development Fund “A way to make Europe”) and the Centro de Investigación Biomédica en Red de Enfermedades Raras (CIBERER), an initiative of the Instituto de Salud Carlos III (Ministerio de Ciencia e Innovación, Spain), the Departament de Salut, Generalitat de Catalunya (URDCAT project, SLT002/16/00174), the Agència de Gestió d’Ajuts Universitaris i de Recerca (AGAUR) (2017: SGR 1428) and the CERCA Programme/Generalitat de Catalunya. TK is a member of the European Reference Network for Rare Neurological Diseases (ERN-RND-Project ID No 739510). TBH was supported by the German Bundesministerium für Bildung und Forschung (BMBF) through the Juniorverbund in der Systemmedizin “mitOmics” (FKZ 01ZX1405). The views expressed in this publication are those of the author(s) and not necessarily those of the NHS, the NIHR, or the Department of Health and Social Care.

## References

Barca, E., Long, Y., Cooley, V., Schoenaker, R., Emmanuele, V., Dimauro, S., Cohen, B. H., Karaa, A., Vladutiu, G. D., Haas, R., Van Hove, J. L. K., Scaglia, F., Parikh, S., Bedoyan, J. K., Debrosse, S. D., Gavrilova, R. H., Saneto, R. P., Enns, G. M., Stacpoole, P. W., … Hirano, M. (2020). Mitochondrial diseases in North America: An analysis of the NAMDC Registry. Neurology: Genetics, 6(2). https://doi.org/10.1212/NXG.0000000000000402

Bick, D., Bick, S. L., Dimmock, D. P., Fowler, T. A., Caulfield, M. J., & Scott, R. H. (2021). An online compendium of treatable genetic disorders. American Journal of Medical Genetics Part C: Seminars in Medical Genetics, 187(1), 48–54. https://doi.org/10.1002/ajmg.c.31874

Bonora, E., Chakrabarty, S., Kellaris, G., Tsutsumi, M., Bianco, F., Bergamini, C., … & De Giorgio, R. (2021). Biallelic variants in LIG3 cause a novel mitochondrial neurogastrointestinal encephalomyopathy. Brain.

Distelmaier, F., Huppke, P., Pieperhoff, P., Amunts, K., Schaper, J., Morava, E., … & Karenfort, M. (2013). Biotin-responsive basal ganglia disease: a treatable differential diagnosis of leigh syndrome. In JIMD Reports-Case and Research Reports, Volume 13 (pp. 53–57). Springer, Berlin, Heidelberg.

Distelmaier, F., Haack, T. B., Wortmann, S. B., Mayr, J. A., & Prokisch, H. (2017). Treatable mitochondrial diseases: cofactor metabolism and beyond. Brain, 140(2), e11–e11.

Firth, H. V., Richards, S. M., Bevan, A. P., Clayton, S., Corpas, M., Rajan, D., … & Carter, N. P. (2009). DECIPHER: database of chromosomal imbalance and phenotype in humans using ensembl resources. The American Journal of Human Genetics, 84(4), 524–533.

French, C. E., Delon, I., Dolling, H., Sanchis-Juan, A., Shamardina, O., Mégy, K., … & Raymond, F. L. (2019). Whole genome sequencing reveals that genetic conditions are frequent in intensively ill children. Intensive care medicine, 45(5), 627–636.

Ganetzky, R. D., Stendel, C., McCormick, E. M., Zolkipli-Cunningham, Z., Goldstein, A. C., Klopstock, T., & Falk, M. J. (2019). MT-ATP6 mitochondrial disease variants: Phenotypic and biochemical features analysis in 218 published cases and cohort of 14 new cases. Human Mutation, 40(5), 499–515. https://doi.org/10.1002/humu.23723

Gorman, G. S., Chinnery, P. F., DiMauro, S., Hirano, M., Koga, Y., McFarland, R., Suomalainen, A., Thorburn, D. R., Zeviani, M., & Turnbull, D. M. (2016). Mitochondrial diseases. Nature Reviews Disease Primers, 2. https://doi.org/10.1038/nrdp.2016.80

Gorman, G. S., Schaefer, A. M., Ng, Y., Gomez, N., Blakely, E. L., Alston, C. L., Feeney, C., Horvath, R., Yu-Wai-Man, P., Chinnery, P. F., Taylor, R. W., Turnbull, D. M., & McFarland, R. (2015). Prevalence of nuclear and mitochondrial DNA mutations related to adult mitochondrial disease. Annals of Neurology, 77(5), 753–759. https://doi.org/10.1002/ana.24362

Greene, D., Richardson, S., & Turro, E. (2017). OntologyX: A suite of R packages for working with ontological data. Bioinformatics, 33(7), 1104–1106. https://doi.org/10.1093/bioinformatics/btw763

Guillen Sacoto, M. J., Tchasovnikarova, I. A., Torti, E., Forster, C., Andrew, E. H., Anselm, I., Baranano, K. W., Briere, L. C., Cohen, J. S., Craigen, W. J., Cytrynbaum, C., Ekhilevitch, N., Elrick, M. J., Fatemi, A., Fraser, J. L., Gallagher, R. C., Guerin, A., Haynes, D., High, F. A., … Juusola, J. (2020). De Novo Variants in the ATPase Module of MORC2 Cause a Neurodevelopmental Disorder with Growth Retardation and Variable Craniofacial Dysmorphism. American Journal of Human Genetics, 107(2), 352–363. https://doi.org/10.1016/j.ajhg.2020.06.013

Haack, T. B., Makowski, C., Yao, Y., Graf, E., Hempel, M., Wieland, T., … & Prokisch, H. (2012). Impaired riboflavin transport due to missense mutations in SLC52A2 causes Brown-Vialetto-Van Laere syndrome. Journal of inherited metabolic disease, 35(6), 943–948.

Haack, T. B., Jackson, C. B., Murayama, K., Kremer, L. S., Schaller, A., Kotzaeridou, U., de Vries, M. C., Schottmann, G., Santra, S., Büchner, B., Wieland, T., Graf, E., Freisinger, P., Eggimann, S., Ohtake, A., Okazaki, Y., Kohda, M., Kishita, Y., Tokuzawa, Y., … Klopstock, T. (2015). Deficiency of ECHS1 causes mitochondrial encephalopathy with cardiac involvement. Annals of Clinical and Translational Neurology, 2(5), 492–509. https://doi.org/10.1002/acn3.189

Hoytema van Konijnenburg, E. M. M., Wortmann, S. B., Koelewijn, M. J., Tseng, L. A., Houben, R., Stöckler-Ipsiroglu, S., Ferreira, C. R., & van Karnebeek, C. D. M. (2021). Treatable inherited metabolic disorders causing intellectual disability: 2021 review and digital app. Orphanet Journal of Rare Diseases, 16(1), 170. https://doi.org/10.1186/s13023-021-01727-2

Kohda, M., Tokuzawa, Y., Kishita, Y., Nyuzuki, H., Moriyama, Y., Mizuno, Y., Hirata, T., Yatsuka, Y., Yamashita-Sugahara, Y., Nakachi, Y., Kato, H., Okuda, A., Tamaru, S., Borna, N. N., Banshoya, K., Aigaki, T., Sato-Miyata, Y., Ohnuma, K., Suzuki, T., … Okazaki, Y. (2016). A Comprehensive Genomic Analysis Reveals the Genetic Landscape of Mitochondrial Respiratory Chain Complex Deficiencies. PLOS Genetics, 12(1), e1005679. https://doi.org/10.1371/journal.pgen.1005679

Köhler, S., Gargano, M., Matentzoglu, N., Carmody, L. C., Lewis-Smith, D., Vasilevsky, N. A., Danis, D., Balagura, G., Baynam, G., Brower, A. M., Callahan, T. J., Chute, C. G., Est, J. L., Galer, P. D., Ganesan, S., Griese, M., Haimel, M., Pazmandi, J., Hanauer, M., … Robinson, P. N. (2021). The human phenotype ontology in 2021. Nucleic Acids Research, 49(D1), D1207–D1217. https://doi.org/10.1093/nar/gkaa1043

Kopajtich, R., Smirnov, D., Stenton, S. L., Loipfinger, S., Meng, C., Scheller, I. F., Freisinger, P., Baski, R., Berutti, R., Behr, J., Bucher, M., Distelmaier, F., Gusic, M., Hempel, M., Kulterer, L., Mayr, J., Meitinger, T., Mertes, C., Metodiev, M. D., … Prokisch, H. (2021). Integration of proteomics with genomics and transcriptomics increases the diagnostic rate of Mendelian disorders. MedRxiv.

Kremer, L. S., Bader, D. M., Mertes, C., Kopajtich, R., Pichler, G., Iuso, A., … & Prokisch, H. (2017). Genetic diagnosis of Mendelian disorders via RNA sequencing. Nature communications, 8(1), 1–11.

Legati, A., Reyes, A., Nasca, A., Invernizzi, F., Lamantea, E., Tiranti, V., Garavaglia, B., Lamperti, C., Ardissone, A., Moroni, I., Robinson, A., Ghezzi, D., & Zeviani, M. (2016). New genes and pathomechanisms in mitochondrial disorders unraveled by NGS technologies. Biochimica et Biophysica Acta - Bioenergetics, 1857(8), 1326–1335. https://doi.org/10.1016/j.bbabio.2016.02.022

Lissens, W., De Meirleir, L., Seneca, S., Liebaers, I., Brown, G. K., Brown, R. M., Ito, M., Naito, E., Kuroda, Y., Kerr, D. S., Wexler, I. D., Patel, M. S., Robinson, B. H., Seyda, A., & Lissens, : W. (2000). Mutations in the X-Linked Pyruvate Dehydrogenase (E1) α Subunit Gene (PDHA1) in Patients With a Pyruvate Dehydrogenase Complex Deficiency. HUMAN MUTATION, 15, 209–219. https://doi.org/10.1002/(SICI)1098-1004(200003)15:3

Marti-Sanchez, L., Baide-Mairena, H., Marcé-Grau, A., Pons, R., Skouma, A., López-Laso, E., Sigatullina, M., Rizzo, C., Semeraro, M., Martinelli, D., Carrozzo, R., Dionisi-Vici, C., González-Gutiérrez-Solana, L., Correa-Vela, M., Ortigoza-Escobar, J. D., Sánchez-Montañez, Á., Vazquez, É., Delgado, I., Aguilera-Albesa, S., … Pérez-Dueñas, B. (2020). Delineating the neurological phenotype in children with defects in the ECHS1 or HIBCH gene. Journal of Inherited Metabolic Disease, 44(2). https://doi.org/10.1002/jimd.12288

Morava, E., Van Den Heuvel, L., Hol, F., De Vries, M. C., Hogeveen, M., Rodenburg, R. J., & Smeitink, J. A. M. (2006). Mitochondrial disease criteria: Diagnostic applications in children. Neurology, 67(10), 1823–1826. https://doi.org/10.1212/01.wnl.0000244435.27645.54

Ng, Y. S., Martikainen, M. H., Gorman, G. S., Blain, A., Bugiardini, E., Bunting, A., Schaefer, A. M., Alston, C. L., Blakely, E. L., Sharma, S., Hughes, I., Lim, A., de Goede, C., McEntagart, M., Spinty, S., Horrocks, I., Roberts, M., Woodward, C. E., Chinnery, P. F., … McFarland, R. (2019). Pathogenic variants in MT-ATP6: A United Kingdom–based mitochondrial disease cohort study. Annals of Neurology, 86(2), 310–315. https://doi.org/10.1002/ana.25525

Ogawa, E., Shimura, M., Fushimi, T., Tajika, M., Ichimoto, K., Matsunaga, A., Tsuruoka, T., Ishige, M., Fuchigami, T., Yamazaki, T., Mori, M., Kohda, M., Kishita, Y., Okazaki, Y., Takahashi, S., Ohtake, A., & Murayama, K. (2017). Clinical validity of biochemical and molecular analysis in diagnosing Leigh syndrome: a study of 106 Japanese patients. Journal of Inherited Metabolic Disease, 40(5), 685–693. https://doi.org/10.1007/s10545-017-0042-6

Ohtake, A., Murayama, K., Mori, M., Harashima, H., Yamazaki, T., Tamaru, S., Yamashita, Y., Kishita, Y., Nakachi, Y., Kohda, M., Tokuzawa, Y., Mizuno, Y., Moriyama, Y., Kato, H., & Okazaki, Y. (2014). Diagnosis and molecular basis of mitochondrial respiratory chain disorders: Exome sequencing for disease gene identification. Biochimica et Biophysica Acta - General Subjects, 1840(4), 1355–1359. https://doi.org/10.1016/j.bbagen.2014.01.025

Pronicka, E., Piekutowska-Abramczuk, D., Ciara, E., Trubicka, J., Rokicki, D., Karkucinska-Wieckowska, A., Pajdowska, M., Jurkiewicz, E., Halat, P., Kosinska, J., Pollak, A., Rydzanicz, M., Stawinski, P., Pronicki, M., Krajewska-Walasek, M., & Płoski, R. (2016). New perspective in diagnostics of mitochondrial disorders: Two years’ experience with whole-exome sequencing at a national paediatric centre. Journal of Translational Medicine, 14(1), 174. https://doi.org/10.1186/s12967-016-0930-9

Puusepp, S., Reinson, K., Pajusalu, S., Murumets, Ü., Õiglane-Shlik, E., Rein, R., Talvik, I., Rodenburg, R. J., & Õunap, K. (2018). Effectiveness of whole exome sequencing in unsolved patients with a clinical suspicion of a mitochondrial disorder in Estonia. Molecular Genetics and Metabolism Reports, 15, 80–89. https://doi.org/10.1016/j.ymgmr.2018.03.004

Rahman, S., & Copeland, W. C. (2019). POLG-related disorders and their neurological manifestations. Nature Reviews Neurology, 15(1), 40–52. https://doi.org/10.1038/s41582-018-0101-0

Repp, B. M., Mastantuono, E., Alston, C. L., Schiff, M., Haack, T. B., Rötig, A., Ardissone, A., Lombès, A., Catarino, C. B., Diodato, D., Schottmann, G., Poulton, J., Burlina, A., Jonckheere, A., Munnich, A., Rolinski, B., Ghezzi, D., Rokicki, D., Wellesley, D., … Wortmann, S. (2018). Clinical, biochemical and genetic spectrum of 70 patients with ACAD9 deficiency: is riboflavin supplementation effective? Orphanet Journal of Rare Diseases, 13(1), 120. https://doi.org/10.1186/s13023-018-0784-8

Richards, S., Aziz, N., Bale, S., Bick, D., Das, S., Gastier-Foster, J., Grody, W. W., Hegde, M., Lyon, E., Spector, E., Voelkerding, K., & Rehm, H. L. (2015). Standards and guidelines for the interpretation of sequence variants: A joint consensus recommendation of the American College of Medical Genetics and Genomics and the Association for Molecular Pathology. Genetics in Medicine, 17(5), 405–424. https://doi.org/10.1038/gim.2015.30

Riley, L. G., Cowley, M. J., Gayevskiy, V., Minoche, A. E., Puttick, C., Thorburn, D. R., Rius, R., Compton, A. G., Menezes, M. J., Bhattacharya, K., Coman, D., Ellaway, C., Alexander, I. E., Adams, L., Kava, M., Robinson, J., Sue, C. M., Balasubramaniam, S., & Christodoulou, J. (2020). The diagnostic utility of genome sequencing in a pediatric cohort with suspected mitochondrial disease. Genetics in Medicine, 22(7), 1254–1261. https://doi.org/10.1038/s41436-020-0793-6

Russell, O. M., Gorman, G. S., Lightowlers, R. N., & Turnbull, D. M. (2020). Mitochondrial diseases: hope for the future. Cell, 181(1), 168–188.

Schlieben, L. D., & Prokisch, H. (2020). The Dimensions of Primary Mitochondrial Disorders. Frontiers in Cell and Developmental Biology, 8. https://doi.org/10.3389/fcell.2020.600079

Stendel, C., Neuhofer, C., Floride, E., Yuqing, S., Ganetzky, R. D., Park, J., Freisinger, P., Kornblum, C., Kleinle, S., Schöls, L., Distelmaier, F., Stettner, G. M., Büchner, B., Falk, M. J., Mayr, J. A., Synofzik, M., Abicht, A., Haack, T. B., Prokisch, H., … Klopstock, T. (2020). Delineating MT-ATP6-associated disease: From isolated neuropathy to early onset neurodegeneration. Neurology: Genetics, 6(1), 393. https://doi.org/10.1212/NXG.0000000000000393

Stenton, S. L., & Prokisch, H. (2020). Genetics of mitochondrial diseases: Identifying mutations to help diagnosis. EBioMedicine, 56, 102784. https://doi.org/10.1016/j.ebiom.2020.102784

Stenton, S. L., Sheremet, N. L., Catarino, C. B., Andreeva, N., Assouline, Z., Barboni, P., Barel, O., Berutti, R., Bychkov, I. O., Caporali, L., Capristo, M., Carbonelli, M., Cascavilla, M. L., Charbel Issa, P., Freisinger, P., Gerber, S., Ghezzi, D., Graf, E., Heidler, J., … Prokisch, H. (2021). Impaired complex I repair causes recessive Leber’s hereditary optic neuropathy. Journal of Clinical Investigation. https://doi.org/10.1172/jci138267

Tan, J., Wagner, M., Stenton, S. L., Strom, T. M., Wortmann, S. B., Prokisch, H., Meitinger, T., Oexle, K., & Klopstock, T. (2020). Lifetime risk of autosomal recessive mitochondrial disorders calculated from genetic databases. EBioMedicine, 54. https://doi.org/10.1016/j.ebiom.2020.102730

Taylor, R. W., Pyle, A., Griffin, H., Blakely, E. L., Duff, J., He, L., Smertenko, T., Alston, C. L., Neeve, V. C., Best, A., Yarham, J. W., Kirschner, J., Schara, U., Talim, B., Topaloglu, H., Baric, I., Holinski-Feder, E., Abicht, A., Czermin, B., … Chinnery, P. F. (2014). Use of whole-exome sequencing to determine the genetic basis of multiple mitochondrial respiratory chain complex deficiencies. JAMA - Journal of the American Medical Association, 312(1), 68–77. https://doi.org/10.1001/jama.2014.7184

Tetreault, M., Fahiminiya, S., Antonicka, H., Mitchell, G. A., Geraghty, M. T., Lines, M., Boycott, K. M., Shoubridge, E. A., Mitchell, J. J., Michaud, J. L., & Majewski, J. (2015). Whole-exome sequencing identifies novel ECHS1 mutations in Leigh syndrome. Human Genetics, 134(9), 981–991. https://doi.org/10.1007/s00439-015-1577-y

Theunissen, T. E. J., Nguyen, M., Kamps, R., Hendrickx, A. T., Sallevelt, S. C. E. H., Gottschalk, R. W. H., Calis, C. M., Stassen, A. P. M., De Koning, B., Mulder-Den Hartog, E. N. M., Schoonderwoerd, K., Fuchs, S. A., Hilhorst-Hofstee, Y., De Visser, M., Vanoevelen, J., Szklarczyk, R., Gerards, M., De Coo, I. F. M., Hellebrekers, D. M. E. I., & Smeets, H. J. M. (2018). Whole exome sequencing is the preferred strategy to identify the genetic defect in patients with a probable or possible mitochondrial cause. Frontiers in Genetics, 9(OCT), 400. https://doi.org/10.3389/fgene.2018.00400

Witters, P., Saada, A., Honzik, T., Tesarova, M., Kleinle, S., Horvath, R., Goldstein, A., & Morava, E. (2018). Revisiting mitochondrial diagnostic criteria in the new era of genomics. Genetics in Medicine, 20(4), 444–451. https://doi.org/10.1038/gim.2017.125

Wortmann, S. B., Koolen, D. A., Smeitink, J. A., van den Heuvel, L., & Rodenburg, R. J. (2015). Whole exome sequencing of suspected mitochondrial patients in clinical practice. Journal of Inherited Metabolic Disease, 38(3), 437–443. https://doi.org/10.1007/s10545-015-9823-y

Wortmann, S. B., Mayr, J. A., Nuoffer, J. M., Prokisch, H., & Sperl, W. (2017). A Guideline for the Diagnosis of Pediatric Mitochondrial Disease: The Value of Muscle and Skin Biopsies in the Genetics Era. Neuropediatrics, 48(4), 309–314. https://doi.org/10.1055/s-0037-1603776

Yépez, V. A., Gusic, M., Kopajtich, R., Mertes, C., Smith, N. H., Alston, C. L., Berutti, R., Blessing, H., Ciara, E., Procopio, E., Ribes, A., Rötig, A., Schwarzmayr, T., Staufner, C., Stenton, S. L., Strom, T. M., Taylor, R. W., Terrile, C., Tort, F., … Prokisch, H. (2021). Clinical implementation of RNA sequencing for Mendelian disease diagnostics. Dorota Piekutowska-Abramczuk, 10, 2021.04.01.21254633. https://doi.org/10.1101/2021.04.01.21254633

Zhou, J., Li, J., Stenton, S. L., Ren, X., Gong, S., Fang, F., & Prokisch, H. (2020). NAD (P) HX dehydratase (NAXD) deficiency: a novel neurodegenerative disorder exacerbated by febrile illnesses. Brain, 143(2), e8–e8.

Zhou, L., Deng, J., Stenton, S. L., Zhou, J., Li, H., Chen, C., … & Fang, F. (2020). Case Report: Rapid Treatment of Uridine-Responsive Epileptic Encephalopathy Caused by CAD Deficiency. Frontiers in Pharmacology, 11, 1956.

