## Supplemental Material for "Diagnosing pediatric mitochondrial disease: lessons from 2,000 exomes"

### Supplemental Figures

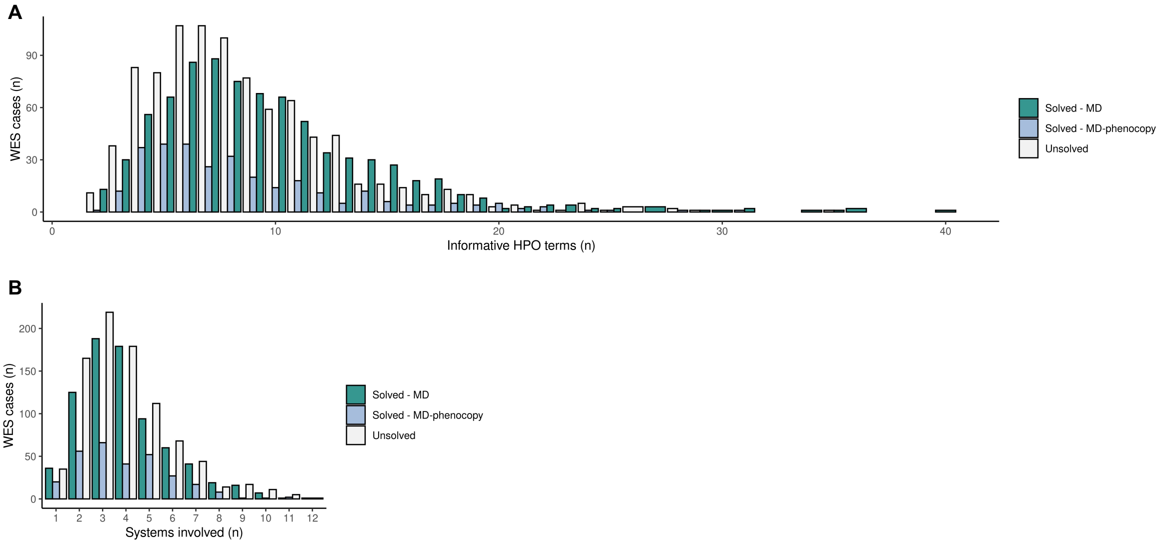


**Supplemental Fig. 1. Number of HPO phenotypes collected per patient. A,** Number of informative (non-redundant) HPO phenotypes collected per patient, stratified by underlying molecular diagnosis. **B,** Number of organ systems involved in the patient's clinical presentation, stratified by underlying molecular diagnosis.

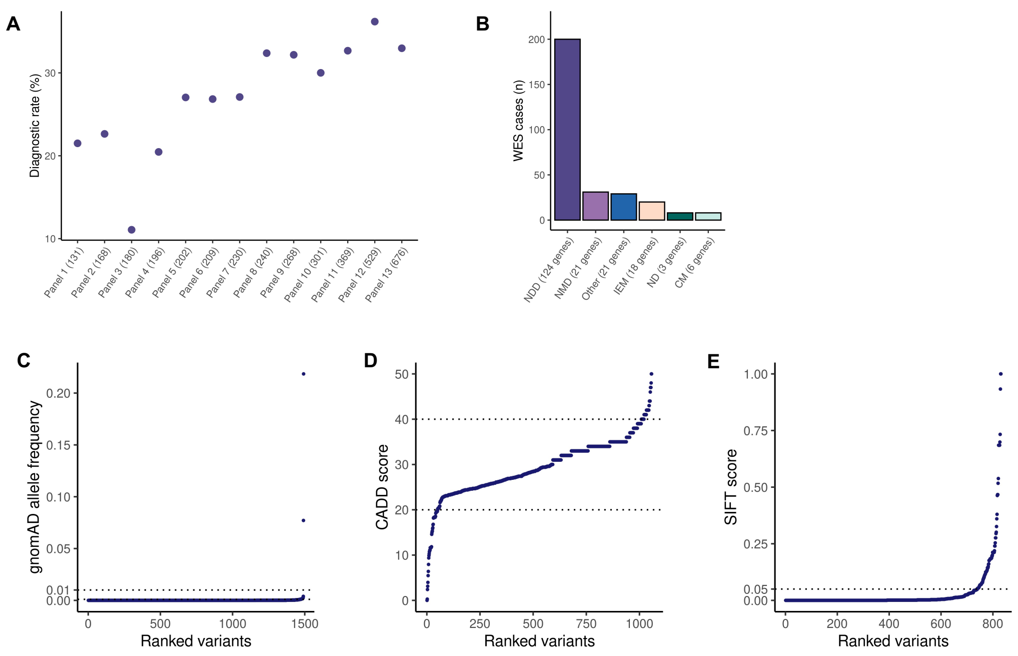


**Supplemental Fig. 2. Molecular etiology of the MDs and MD-phenocopies. A,** Theoretical diagnostic rate achievable in the study cohort by the application of 13 different MD panels (see **Supplementary Methods** for details). **B,** Number of MD-phenocopy cases by disease gene category. **C,** All identified disease-causing variants ranked by gnomAD allele frequency. Routinely applied allele frequency thresholds of 1% and 0.1% are indicated. Two frequent variants in *DARS2* were confirmed to be pathogenic as a rare combination in *cis*, confirmed by RNA seq and proteomic analyses as reported by Kopajtich et al., 2021. **D,** All identified disease-causing variants ranked by CADD score. CADD scores ≥20 and ≥40 are indicated. Variants above these thresholds are within the 1% and 0.1% most deleterious substitutions, respectively. **E,** All identified disease-causing variants ranked by SIFT score. respectively. SIFT scores <0.05 are predicted to be deleterious. NDD, neurodevelopmental disease; NMD, neuromuscular disease; IEM, inborn errors of metabolism; ND, neurodegeneration; CM, cardiomyopathy.

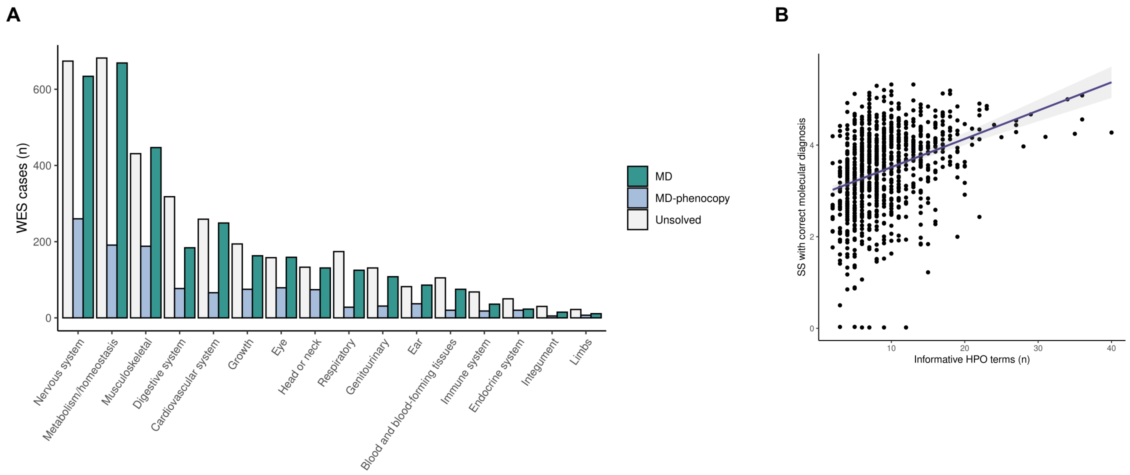


**Supplemental Fig. 3. Association between phenotypic presentation and molecular diagnosis. A,** Frequency of organ system involvement, stratified by underlying molecular diagnosis. **B,** Correlation between the number of informative (non-redundant) HPO terms (reflecting more complex phenotypes) and the semantic similarity (SS) of the correct molecular diagnosis amongst all OMIM disease genes based on patient HPO terms.

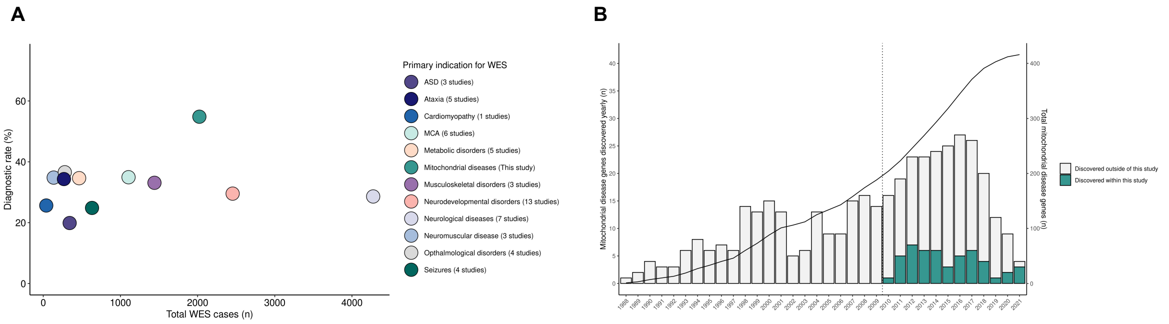


**Supplemental Fig. 4. WES diagnostic rate and MD-associated gene overview. A,** The percentage of patients investigated by WES receiving a genetic diagnosis indicated for a variety of primary metabolic and neurological indications. Studies were included based on the disease under investigation and whether exome sequencing was undertaken in the context of a diagnostic test (Schabhüttl et al., 2014, Iglesias et al., 2014, Lee et al., 2014, Sawyer et al., 2014, Pyle et al., 2015, Farwell et al., 2015, Zhu et al., 2015, Sawyer et al., 2016, Tarailo-Graovac et al., 2016, Thevenon et al., 2016, Retterer et al., 2016, Trujillano et al., 2017, Theunissen et al., 2018). The total number of patients sequenced and total number of studies included is indicated. **B,** Discovery of MD-associated genes between 1988 and 2021. The vertical line indicates transition from mtDNA and candidate gene sequencing to next-generation sequencing (NGS) in 2010, reflected by an acceleration in disease gene discovery. In total, 416 MD-associated genes have been described to date of which 49 were discovered within this study cohort. ASD, autism spectrum disorders; MCA, multiple-congenital abnormalities.

### Supplementary Methods

#### Whole exome sequencing (WES) analysis

WES data were collected through international collaboration, initiated by the European Network for Mitochondrial Diseases (GENOMIT). Experimental procedures were performed in the respective centers, as detailed separately in (Taylor et al., 2014, Pronicka et al., 2016, Ruzzenente et al., 2018, Zech at al., 2020, Tort et al., 2020). Variant calling and annotation incorporated numerous publicly available bioinformatics tools and customized software. Reads were aligned to the human reference genome (UCSC Genome Browser build hg19) using Burrows-Wheeler Aligner (v.0.7.5a) (Li and Durbin 2009). Single-nucleotide variants and small insertions and deletions (indels) were detected with SAMtools (v.0.1.19) (Li et al., 2009) and the Genome Analysis Toolkit (GATK) (Van der Auwera et al., 2013). Copy number variants (CNV) were identified with ExomeDepth (Plagnol et al., 2012). Runs of homozygosity (RoH) were detected by BCFtools/RoH, (Narasimhan et al., 2016). As per ACMG guidelines, consanguinity is suspected when RoH accounted for ≥10% of the genome, consistent with a first- or second-degree parental relationship (Rehder et al., 2013). Mitochondrial genome (mtDNA) variants were called from the exome data as described in Wagner et al., 2019. The reported variants were annotated with their allele frequency (gnomAD database, http://gnomad.broadinstitute.org/), and predicted functional consequence on the gene product. Variants were prioritized by predicted deleterious effects on the protein by the Combined Annotation Dependent Depletion (CADD) and Sorting Intolerance from Tolerance (SIFT) scores. Variants prioritized as potentially disease-causing were formally classified by ACMG recommendations (Richards et al., 2015) using the Python package 'InterVar' (Li and Wang 2017) and were subjected to co-segregation analyses. Semantic similarity ≥2 was accepted as a phenotype match (ACMG criteria PP4) (as per Kopajtich et al., 2021). A mitochondrial RCC defect on tissue biopsy or a maximal respiration rate <71.6% in patient-derived fibroblast cell lines (Ogawa et al., 2017) were accepted as functional evidence for variants in MD genes (ACMG criteria PS3). In autosomal and X-linked dominant disease genes, a pathogenic variant (P, ACMG class 5) or a likely pathogenic variant (LP, class 4) was reported as a definite molecular diagnosis. In autosomal recessive and X-linked disease genes, biallelic or hemizygous P/LP variants were reported as definite molecular diagnoses. Patients with mtDNA depletion or multiple mtDNA deletions and no identified causative variant(s) were not considered to be solved.

**“Mitochondrial disease” panel selection for calculation of theoretical diagnostic rate**

We searched the Genetic Testing Registry (GTR) of NCBI (accessed June 2020) for providers of commercial panels for “mitochondrial disease” identifying 10 diagnostic panels of between 100 to 1000 genes and added three additional custom diagnostic panels used by centers in the study not registered at GTR, to in total evaluate 13 diagnostic panels targeting MD, as listed below. The selected panels covered a total of 1,099 different disease and candidate disease genes (mean 285 genes ± 157 genes s.d. per panel). Gene lists are freely available for download from the respective providers’ websites or available upon request, as indicated.

| Panel identifier | Panel provider | Panel size |
|---|---|---|
| Mitochondrial Encephalopathy | MGZ Medical Genetics Center | 131 |
| Mitochondrial Diseases | MGZ Medical Genetics Center | 168 |
| Comprehensive mitochondrial disorders panel | Centogene AG - the Rare Disease Company | 180 |
| Comprehensive Mitochondrial Metabolic Panel | Knight Diagnostic Laboratories - Molecular Diagnostic Center | 196 |
| Mitochondrial Focused Nuclear Gene Panel | GeneDx | 202 |
| Mitochondrial Diseases | Asper Biogene | 209 |
| MitONE230 | Upon request* | 230 |
| Combined Mito Genome Plus Mito Focused Nuclear Gene Panel | GeneDx | 240 |
| Mitochondrial diseases | CGC Genetics | 268 |
| MitoSure300 | Upon request* | 300 |
| WES mitochondrial disorders | Translational Metabolic Laboratory | 369 |
| Metabolic Diseases incl. Mitochondriopathies | CeGaT | 529 |
| Nuclear-Mito NGS Panel | Fulgent Genetics | 676 |

*Custom panels utilised by the Unit of Medical Genetics and Neurogenetics, Fondazione IRCCS Istituto Neurologico Carlo Besta, contacts:

### Modified Nijmegen Mitochondrial Disease Criteria standardized for HPO term usage

| **Clinical signs and symptoms, 1 point/symptom (max. 4 points)** | | | **Metabolic/ imaging studies (max. 4 points)** | **Tissue histopathology and biochemistry (max. 4 points)** |
|---|---|---|---|---|
| Muscular presentation (max. 2 points) | CNS presentation (max. 2 points) | Multisystem disease (max. 3 points) | | |
| Ophthalmoplegia (HP:0000602)* | Neurodevelopmental abnormality (HP:0012759) **or** Intellectual disability (HP:0001249) | Hematological abnormality (HP:0001871) **or** Immune abnormality (HP:0002715) | Increased serum lactate (HP:0002151)* | Ragged-red muscle fibers (HP:0003200)** |
| Ptosis (HP:0000508) **or** Myopathic facies (HP:0002058) | Delayed speech and language development (HP:0000750) | (Digestive system abnormality (HP:0025031) **or** Decreased liver function (HP:0001410)* **or** Abnormality of the liver (HP:0001392)* | Elevated lactate:pyruvate ratio (HP:0032653) | COX negative muscle fibers (HP:0003688)** |
| Exercise intolerance (HP:0003546) **or** Fatigue (HP:0012378) | Developmental regression (HP:0002376) | Endocrine abnormality (HP:0000818) | Increased serum alanine (HP:0003348)* | Abnormal mitochondrial morphology (HP:0008322)* |
| Muscle weakness (HP:0001324) **or** Myopathy (HP:0003198) **or** Muscular hypotonia (HP:0001252) | Stroke-like episode (HP:0002401) | Growth abnormality (HP:0001507) | Increased CSF lactate (HP:0002490)* | Decreased activity of mitochondrial respiratory chain (HP:0008972)* **or** Decreased activity of the pyruvate dehydrogenase complex (HP:0002928)* |
| Rhabdomyolsis (HP:0003201) **or** Increased serum creatine kinase (HP:0003236) | Migraine (HP:0002076) | Abnormality of the cardiovascular system (HP:0001626) **or** Cardiomyopathy (HP:0001638)* | Increased CSF alanine (HP:0500233)* | Mitochondrial depletion (HP:0030059) |
| Motor developmental delay (HP:0001270) | Seizure (HP:0001250) **or** Encepahlopathy (HP:0001298) | Abnormal renal physiology (HP:0012211) | Increased CSF protein (HP:0002922) | |
| EMG abnormality (HP:0003457) | Myoclonus (HP:0001336) | Abnormality of the eye (HP:0000478) **or** Visual impairment (HP:0000505) **or** Optic atrophy (HP:0000648)* **or** Leber optic atrophy (HP:0001112)* | Ethylmalonic aciduria (HP:0003219) **or** Methylmalonic aciduria (HP:0012120) | |
| | Cerebral visual impairment (HP:0100704) | Hearing impairment* (HP:0000365) | 3-Methylglutaconic aciduria (HP:0003535) | |
| | Pyramidal signs (HP:0002493) **or** Spasticity (HP:0001257) | Peripheral neuropathy (HP:0009830) | Stroke-like picture (HP:0002401) | |
| | Extrapyramidal signs (HP:0002071) **or** Dystonia (HP:0001332) | Family history (HP:0032316) | Abnormality of the basal ganglia (HP:0002134)* | |
| | Ataxia (HP:0001251) | | Abnormality of the brain stem (HP:0002363)* | |
| | | | Abnormality of the cerebral white matter (HP:0002500) | |
| | | | Elevated brain lactate level by MRS (HP:0012707) | |

*These specific symptoms score 2 points
** These specific symptoms score 4 points

Score 1: unlikely; Score 2-4: possible; Score 5-7: probable; Score 8-12: definite mitochondrial disease

### Modified Revised Mitochondrial Disease Criteria standardized for HPO term usage

| Clinical score, 1 point/symptom (max. 4 points) | | | Metabolic and MRI score, 1 point/symptom (max. 4 points) | |
|---|---|---|---|---|
| Muscular (max. 2 points) | Neurological (max. 2 points) | Multisystem (max. 3 points) | Metabolic (max. 4 points) | Imaging (max. 4 points) |
| Muscle weakness (HP:0001324) **or** Myopathy (HP:0003198) **or** Muscular hypotonia (HP:0001252)<br><br>EMG abnormality (HP:0003457)<br><br>Motor delay (HP:0001270)<br><br>Exercise intolerance (HP:0003546)<br><br>Ophthalmoplegia (HP:0000602) | Neurodevelopmental abnormality (HP:0012759) **or** Intellectual disability (HP:0001249)<br><br>Delayed speech and language development (HP:0000750)<br><br>Dystonia (HP:0001332)<br><br>Ataxia (HP:0001251)<br><br>Spasticity (HP:0001257)<br><br>Peripheral neuropathy (HP:0009830)<br><br>Seizure (HP:0001250) **or** Encephalopathy (HP:0001298) | Digestive system abnormality (HP:0025031)<br><br>Growth delay (HP:0001510) **or** Failure to thrive (HP:0001508)<br><br>Endocrine abnormality (HP:0000818)<br><br>Immune abnormality (HP:0002715)<br><br>Visual impairment (HP:0000505) **or** Hearing impairment (HP:0000365)<br><br>Renal tubular dysfunction (HP:0000124)<br><br>Abnormality of the cardiovascular system (HP:0001626) | Increased serum lactate (HP:0002151)*<br><br>Increased serum alanine (HP:0003348)<br><br>Ethylmalonic aciduria (HP:0003219) **or** Methylmalonic aciduria (HP:0012120)<br><br>3-Methylglutaconic aciduria (HP:0003535)<br><br>Increased CSF lactate (HP:0002490) **or** Increased CSF alanine (HP:0500233) | Abnormality of the basal ganglia (HP:0002134)* **or** Abnormality of the brainstem (HP:0002363)*<br><br>Stroke-like episode (HP:0002401)*<br><br>Elevated brain lactate level by MRS (HP:0012707)<br><br>Abnormality of the cerebral white matter (HP:0002500)<br><br>Abnormality of the thalamus (HP:0010663)<br><br>Agenesis of the corpus callosum (HP:0001274) |

*These specific symptoms score 2 points

Score 1: unlikely; Score 2-4: possible; Score 5-7: probable; Score 8: definite mitochondrial disease

**Supplemental References**


Farwell, K. D., Shahmirzadi, L., El-Khechen, D., Powis, Z., Chao, E. C., Tippin Davis, B., Baxter, R. M., Zeng, W., Mroske, C., Parra, M. C., Gandomi, S. K., Lu, I., Li, X., Lu, H., Lu, H. M., Salvador, D., Ruble, D., Lao, M., Fischbach, S., … Tang, S. (2015). Enhanced utility of family-centered diagnostic exome sequencing with inheritance model-based analysis: Results from 500 unselected families with undiagnosed genetic conditions. Genetics in Medicine, 17(7), 578–586. https://doi.org/10.1038/gim.2014.154

Iglesias, A., Anyane-Yeboa, K., Wynn, J., Wilson, A., Truitt Cho, M., Guzman, E., Sisson, R., Egan, C., & Chung, W. K. (2014). The usefulness of whole-exome sequencing in routine clinical practice. Genetics in Medicine, 16(12), 922–931. https://doi.org/10.1038/gim.2014.58

Kopajtich, R., Smirnov, D., Stenton, S. L., Loipfinger, S., Meng, C., Scheller, I. F., Freisinger, P., Baski, R., Berutti, R., Behr, J., Bucher, M., Distelmaier, F., Gusic, M., Hempel, M., Kulterer, L., Mayr, J., Meitinger, T., Mertes, C., Metodiev, M. D., … Prokisch, H. (2021). Integration of proteomics with genomics and transcriptomics increases the diagnostic rate of Mendelian disorders. MedRxiv.

Lee, H., Deignan, J. L., Dorrani, N., Strom, S. P., Kantarci, S., Quintero-Rivera, F., Das, K., Toy, T., Harry, B., Yourshaw, M., Fox, M., Fogel, B. L., Martinez-Agosto, J. A., Wong, D. A., Chang, V. Y., Shieh, P. B., Palmer, C. G. S., Dipple, K. M., Grody, W. W., … Nelson, S. F. (2014). Clinical exome sequencing for genetic identification of rare mendelian disorders. JAMA - Journal of the American Medical Association, 312(18), 1880–1887. https://doi.org/10.1001/jama.2014.14604

Li, H., & Durbin, R. (2009). Fast and accurate short read alignment with Burrows-Wheeler transform. Bioinformatics, 25(14), 1754–1760. https://doi.org/10.1093/bioinformatics/btp324

Li, H., Handsaker, B., Wysoker, A., Fennell, T., Ruan, J., Homer, N., Marth, G., Abecasis, G., & Durbin, R. (2009). The Sequence Alignment/Map format and SAMtools. Bioinformatics, 25(16), 2078–2079. https://doi.org/10.1093/bioinformatics/btp352

Li, Q., & Wang, K. (2017). InterVar: Clinical Interpretation of Genetic Variants by the 2015 ACMG-AMP Guidelines. American Journal of Human Genetics, 100(2), 267–280. https://doi.org/10.1016/j.ajhg.2017.01.004

Narasimhan, V., Danecek, P., Scally, A., Xue, Y., Tyler-Smith, C., & Durbin, R. (2016). BCFtools/RoH: A hidden Markov model approach for detecting autozygosity from next-generation sequencing data. Bioinformatics, 32(11), 1749–1751. https://doi.org/10.1093/bioinformatics/btw044

Ogawa, E., Shimura, M., Fushimi, T., Tajika, M., Ichimoto, K., Matsunaga, A., Tsuruoka, T., Ishige, M., Fuchigami, T., Yamazaki, T., Mori, M., Kohda, M., Kishita, Y., Okazaki, Y., Takahashi, S., Ohtake, A., & Murayama, K. (2017). Clinical validity of biochemical and molecular analysis in diagnosing Leigh syndrome: a study of 106 Japanese patients. Journal of Inherited Metabolic Disease, 40(5), 685–693.

Plagnol, V., Curtis, J., Epstein, M., Mok, K. Y., Stebbings, E., Grigoriadou, S., Wood, N. W., Hambleton, S., Burns, S. O., Thrasher, A. J., Kumararatne, D., Doffinger, R., & Nejentsev, S. (2012). A robust model for read count data in exome sequencing experiments and implications for copy number variant calling. Bioinformatics, 28(21), 2747–2754. https://doi.org/10.1093/bioinformatics/bts526

Pronicka, E., Piekutowska-Abramczuk, D., Ciara, E., Trubicka, J., Rokicki, D., Karkucinska-Wieckowska, A., Pajdowska, M., Jurkiewicz, E., Halat, P., Kosinska, J., Pollak, A., Rydzanicz, M., Stawinski, P., Pronicki, M., Krajewska-Walasek, M., & Płoski, R. (2016). New perspective in diagnostics of mitochondrial disorders: Two years' experience with whole-exome sequencing at a national paediatric centre. Journal of Translational Medicine, 14(1), 174. https://doi.org/10.1186/s12967-016-0930-9

Pyle, A., Smertenko, T., Bargiela, D., Griffin, H., Duff, J., Appleton, M., Douroudis, K., Pfeffer, G., Santibanez-Koref, M., Eglon, G., Yu-Wai-Man, P., Ramesh, V., Horvath, R., & Chinnery, P. F. (2015). Exome sequencing in undiagnosed inherited and sporadic ataxias. Brain, 138(2), 276–283. https://doi.org/10.1093/brain/awu348

Rehder, C. W., David, K. L., Hirsch, B., Toriello, H. V., Wilson, C. M., & Kearney, H. M. (2013). American College of medical genetics and genomics: Standards and guidelines for documenting suspected consanguinity as an incidental finding of genomic testing. Genetics in Medicine, 15(2), 150–152. https://doi.org/10.1038/gim.2012.169

Retterer, K., Juusola, J., Cho, M. T., Vitazka, P., Millan, F., Gibellini, F., Vertino-bell, A., Smaoui, N., Neidich, J., Monaghan, K. G., Mcknight, D., Bai, R., Suchy, S., Friedman, B., Tahiliani, J., Pineda-alvarez, D., Richard, G., Brandt, T., Haverfield, E., … Bale, S. (2016). Clinical application of whole-exome sequencing across clinical indications. 18(7). https://doi.org/10.1038/gim.2015.148

Richards, S., Aziz, N., Bale, S., Bick, D., Das, S., Gastier-Foster, J., Grody, W. W., Hegde, M., Lyon, E., Spector, E., Voelkerding, K., & Rehm, H. L. (2015). Standards and guidelines for the interpretation of sequence variants: A joint consensus recommendation of the American College of Medical Genetics and Genomics and the Association for Molecular Pathology. Genetics in Medicine, 17(5), 405–424. https://doi.org/10.1038/gim.2015.30

Ruzzenente, B., Assouline, Z., Barcia, G., Rio, M., Boddaert, N., Munnich, A., Rötig, A., & Metodiev, M. D. (2018). Inhibition of mitochondrial translation in fibroblasts from a patient expressing the KARS p.(Pro228Leu) variant and presenting with sensorineural deafness, developmental delay, and lactic acidosis. Human Mutation, 39(12), 2047–2059. https://doi.org/10.1002/humu.23657

Sawyer, S. L., Hartley, T., Dyment, D. A., Beaulieu, C. L., Schwartzentruber, J., Smith, A., Bedford, H. M., Bernard, G., Bernier, F. P., Brais, B., Bulman, D. E., Warman Chardon, J., Chitayat, D., Deladoëy, J., Fernandez, B. A., Frosk, P., Geraghty, M. T., Gerull, B., Gibson, W., … Boycott, K. M. (2016). Utility of whole-exome sequencing for those near the end of the diagnostic odyssey: Time to address gaps in care. Clinical Genetics, 89(3), 275–284. https://doi.org/10.1111/cge.12654

Sawyer, Sarah L., Schwartzentruber, J., Beaulieu, C. L., Dyment, D., Smith, A., Chardon, J. W., Yoon, G., Rouleau, G. A., Suchowersky, O., Siu, V., Murphy, L., Hegele, R. A., Marshall, C. R., Bulman, D. E., Majewski, J., Tarnopolsky, M., & Boycott, K. M. (2014). Exome Sequencing as a Diagnostic Tool for Pediatric-Onset Ataxia. Human Mutation, 35(1), 45–49. https://doi.org/10.1002/humu.22451

Schabhüttl, M., Wieland, T., Senderek, J., Baets, J., Timmerman, V., De Jonghe, P., Reilly, M. M., Stieglbauer, K., Laich, E., Windhager, R., Erwa, W., Trajanoski, S., Strom, T. M., & Auer-Grumbach, M. (2014). Whole-exome sequencing in patients with inherited neuropathies: Outcome and challenges. Journal of Neurology, 261(5), 970–982. https://doi.org/10.1007/s00415-014-7289-8

Tarailo-Graovac, M., Shyr, C., Ross, C. J., Horvath, G. A., Salvarinova, R., Ye, X. C., Zhang, L.-H., Bhavsar, A. P., Lee, J. J. Y., Drögemöller, B. I., Abdelsayed, M., Alfadhel, M., Armstrong, L., Baumgartner, M. R., Burda, P., Connolly, M. B., Cameron, J., Demos, M., Dewan, T., … van Karnebeek, C. D. (2016). Exome Sequencing and the Management of Neurometabolic Disorders. New England Journal of Medicine, 374(23), 2246–2255. https://doi.org/10.1056/nejmoa1515792

Taylor, R. W., Pyle, A., Griffin, H., Blakely, E. L., Duff, J., He, L., Smertenko, T., Alston, C. L., Neeve, V. C., Best, A., Yarham, J. W., Kirschner, J., Schara, U., Talim, B., Topaloglu, H., Baric, I., Holinski-Feder, E., Abicht, A., Czermin, B., … Chinnery, P. F. (2014). Use of whole-exome sequencing to determine the genetic basis of multiple mitochondrial respiratory chain complex deficiencies. JAMA - Journal of the American Medical Association, 312(1), 68–77. https://doi.org/10.1001/jama.2014.7184

Theunissen, T. E. J., Nguyen, M., Kamps, R., Hendrickx, A. T., Sallevelt, S. C. E. H., Gottschalk, R. W. H., Calis, C. M., Stassen, A. P. M., De Koning, B., Mulder-Den Hartog, E. N. M., Schoonderwoerd, K., Fuchs, S. A., Hilhorst-Hofstee, Y., De Visser, M., Vanoevelen, J., Szklarczyk, R., Gerards, M., De Coo, I. F. M., Hellebrekers, D. M. E. I., & Smeets, H. J. M. (2018). Whole exome sequencing is the preferred strategy to identify the genetic defect in patients with a probable or possible mitochondrial cause. Frontiers in Genetics, 9(OCT), 400. https://doi.org/10.3389/fgene.2018.00400

Thevenon, J., Duffourd, Y., Masurel-Paulet, A., Lefebvre, M., Feillet, F., El Chehadeh-Djebbar, S., St-Onge, J., Steinmetz, A., Huet, F., Chouchane, M., Darmency-Stamboul, V., Callier, P., Thauvin-Robinet, C., Faivre, L., & Rivière, J. B. (2016). Diagnostic odyssey in severe neurodevelopmental disorders: Toward clinical whole-exome sequencing as a first-line diagnostic test. Clinical Genetics, 89(6), 700–707. https://doi.org/10.1111/cge.12732

Tort, F., Barredo, E., Parthasarathy, R., Ugarteburu, O., Ferrer-Cortès, X., García-Villoria, J., Gort, L., González-Quintana, A., Martín, M. A., Fernández-Vizarra, E., Zeviani, M., & Ribes, A. (2020). Biallelic mutations in NDUFA8 cause complex I deficiency in two siblings with favorable clinical evolution. Molecular Genetics and Metabolism, 131(3), 349–357. https://doi.org/10.1016/j.ymgme.2020.10.005

Trujillano, D., Bertoli-Avella, A. M., Kumar Kandaswamy, K., Weiss, M. E., Köster, J., Marais, A., Paknia, O., Schröder, R., Garcia-Aznar, J. M., Werber, M., Brandau, O., Calvo Del Castillo, M., Baldi, C., Wessel, K., Kishore, S., Nahavandi, N., Eyaid, W., Al Rifai, M. T., Al-Rumayyan, A., … Abou Jamra, R. (2017). Clinical exome sequencing: Results from 2819 samples reflecting 1000 families. European Journal of Human Genetics, 25(2), 176–182. https://doi.org/10.1038/ejhg.2016.146

Van der Auwera, G. A., Carneiro, M. O., Hartl, C., Poplin, R., del Angel, G., Levy-Moonshine, A., Jordan, T., Shakir, K., Roazen, D., Thibault, J., Banks, E., Garimella, K. V., Altshuler, D., Gabriel, S., & DePristo, M. A. (2013). From fastQ data to high-confidence variant calls: The genome analysis toolkit best practices pipeline. Current Protocols in Bioinformatics, 43(SUPL.43). https://doi.org/10.1002/0471250953.bi1110s43

Wagner, M., Berutti, R., Lorenz-Depiereux, B., Graf, E., Eckstein, G., Mayr, J. A., Meitinger, T., Ahting, U., Prokisch, H., Strom, T. M., & Wortmann, S. B. (2019). Mitochondrial DNA mutation analysis from exome sequencing—A more holistic approach in diagnostics of suspected mitochondrial disease. Journal of Inherited Metabolic Disease, 42(5), 909–917. https://doi.org/10.1002/jimd.12109

Zech, M., Jech, R., Boesch, S., Škorvánek, M., Weber, S., Wagner, M., Zhao, C., Jochim, A., Necpál, J., Dincer, Y., Vill, K., Distelmaier, F., Stoklosa, M., Krenn, M., Grunwald, S., Bock-Bierbaum, T., Fečíková, A., Havránková, P., Roth, J., … Winkelmann, J. (2020). Monogenic variants in dystonia: an exome-wide sequencing study. The Lancet Neurology, 19(11), 908–918. https://doi.org/10.1016/S1474-4422(20)30312-4

Zhu, X., Petrovski, S., Xie, P., Ruzzo, E. K., Lu, Y. F., McSweeney, K. M., Ben-Zeev, B., Nissenkorn, A., Anikster, Y., Oz-Levi, D., Dhindsa, R. S., Hitomi, Y., Schoch, K., Spillmann, R. C., Heimer, G., Marek-Yagel, D., Tzadok, M., Han, Y., Worley, G., … Goldstein, D. B. (2015). Whole-exome sequencing in undiagnosed genetic diseases: Interpreting 119 trios. Genetics in Medicine, 17(10), 774–781. https://doi.org/10.1038/gim.2014.191
